# The magnitude of early hepatitis B RNA and DNA declines directly inform capsid assembly modulator effectiveness

**DOI:** 10.64898/2026.08.14.26360479

**Authors:** Tyler Cassidy, Sarafa A. Iyaniwura, Ruy M. Ribeiro, Alan S. Perelson

## Abstract

Capsid assembly modulators (CAMs) are a promising class of antiviral treatments for hepatitis B virus (HBV) infection. Several CAMs have been evaluated in clinical trials but there is no simple method to estimate their *in vivo* antiviral effectiveness. We performed viral dynamics modeling of the intracellular and extracellular dynamics of HBV RNA, HBV DNA, and ALT during phase I trials of two CAMs, vebicorvir and ABI-H2158, which inhibit the encapsidation of pgRNA. Fitting our model to the data, we quantify the drug-induced percent inhibition of encapsidated pgRNA production, which we term their *in vivo* antiviral effectiveness. In both trials, the HBV RNA and HBV DNA declined in a biphasic manner during therapy. The model described these decays well and, by fitting the model to the data, we estimated the CAM effectiveness in each trial participant. Mathematical analysis of the model showed that the magnitude of the first phase of decline of HBV RNA and HBV DNA is explicitly related to CAM effectiveness. However, in the clinic, the end of the first phase may not be known due to sparse sampling. Using clinical trial simulations, we show that the HBV RNA and HBV DNA declines between baseline and day 14 of CAM monotherapy can be used to predict CAM effectiveness. We show that HBV RNA is a clinically relevant biomarker and that very short-term phase I clinical trials can be used to evaluate the in vivo effectiveness of new CAMs, thus reducing the danger of drug resistance developing in trial participants.

## Introduction

Despite the availability of an effective vaccine, approximately 250 million people live with chronic HBV infections (1). These individuals are at increased risk of both liver cirrhosis and hepatocellular carcinoma with chronic HBV infection estimated to be responsible for over 1.1 million deaths per year (1). Existing antiviral therapies, such as nucleoside analogues (NAs), effectively inhibit viral production and slow disease progression. However, existing treatments have low functional cure rates, defined as the sustained loss of HBsAg and HBV DNA following finite treatment (2, 3). In most individuals, viral rebound occurs following treatment interruption. Therefore, chronic HBV infection can require lifelong treatment to control liver damage, and there is a pressing need for the development of new therapies to drive functional cure of chronic HBV.

Capsid assembly modulators (CAMs) are a novel class of direct-acting antivirals that block HBV replication. There are two primary types of CAMs that disrupt viral replication by either inducing the assembly of aberrant capsids (class-A) or stabilizing core protein interactions, leading to the assembly of empty capsids (class-E) (4), although other modes of action have also been implicated (5). The antiviral effects of several CAMs have been evaluated in phase I monotherapy clinical trials, which demonstrated robust declines in both HBV DNA and HBV RNA concentrations (6–10). However, there is no simple method to evaluate the *in vivo* antiviral effectiveness of these CAMs, defined here as the percent inhibition of encapsidated pregenomic RNA (pgRNA) production. Here, we demonstrate that the magnitude of the early HBV RNA and HBV DNA decline directly predict the *in vivo* antiviral efficacy of CAMs and validate our results using data from phase I trials of two first-generation CAMs.

Our modeling links the typical biphasic HBV viral dynamics seen after the start of CAM therapy with the *in vivo* effectiveness of CAMs in blocking viral replication. We predict the time of transition between these phases of decay as a function of CAM effectiveness. We then identify an explicit relationship between the relative declines of HBV RNA and HBV DNA during the first phase and CAM effectiveness, as we previously did for declines of HCV RNA in HCV infection (11, 12). Furthermore, we show that the relative decline of HBV RNA and HBV DNA over 14 days of treatment is sufficient to determine CAM effectiveness. Therefore, our results may facilitate clinical evaluation of the antiviral effects of CAM monotherapy by identifying an explicit relationship between CAM effectiveness and observable viral dynamics. Moreover, CAM resistant HBV variants can arise naturally (13, 14) or emerge by mutagenesis, which resulted in the failure of a recent CAM monotherapy clinical trial (15, 16). Using our proposed method, drug effectiveness can be evaluated with short clinical trials that reduce the risk of drug resistance arising in trial participants, which may preclude them from being treated with CAMs after approval (15, 16).

## Methods

### Viral load data

We considered longitudinal HBV RNA, HBV DNA, and alanine amino transferase (ALT) measurements from a phase 1, randomized, placebo-controlled, multiple ascending dose trial of a first-generation CAM, vebicorvir (NCT02908191) (6). Briefly, 30 participants with chronic HBV infection were randomly assigned to receive 100 mg (n=10), 200 mg (n=10), or 300 mg (n=10) oral doses of vebicorvir once daily (QD) for 28 days. Serum HBV RNA, HBV DNA, and ALT concentrations were measured on days 0 (start of treatment), 1, 7, 14, 21, and 28 during treatment and after treatment cessation on days 35, 42, and 56. The full study inclusion criteria and details are reported elsewhere (6). We excluded one participant with a pre-existing known CAM resistance mutation (Thr109Met) in the 300 mg dose arm. Therefore, in our modeling, we included data from 29 participants in the phase 1 trial of vebicorvir, of whom 17 were HBeAg-positive. The lower limits of detection (LLoD) for HBV DNA and HBV RNA in this trial were 0.95 log_10_ IU/mL and 2.49 log_10_ copies/mL, respectively.

We also considered longitudinal HBV RNA, DNA, and ALT measurements from a phase 1, randomized, placebo-controlled, multiple ascending dose trial of a second CAM-E, ABI-H2158 (NCT03714152) (7). In this trial, 29 HBeAg-positive participants with chronic HBV infection were randomized to receive 100 mg QD (n=7), 300 mg QD (n=7), 500 mg QD (n = 7), or 300 mg twice daily (BID) (n = 8) oral doses of ABI-H2158 for 14 days. HBV RNA, DNA, and ALT concentrations were measured on days 0 (start of treatment), 1, 7, and 13 during treatment, and after treatment cessation on days 14, 21, 28, and 56. One participant each in the 300 mg QD and 300 mg BID cohorts had outlier HBV DNA responses, with significantly lower baseline viral loads and smaller changes from baseline to day 14 than the overall means in their treatment groups. They also exhibited a continued decrease in HBV DNA following treatment cessation (7). One additional participant in the 300 mg BID group was treated with entecavir regularly in the three years preceding the trial and from day 15 onwards (7). These three atypical participants were excluded from our analysis, and we only considered data from the other 26 HBeAg-positive participants. In this trial, the lower limits of quantitation (LLoQ) of HBV DNA and HBV RNA were 1.30 log_10_ IU/mL and 2.4 log_10_ copies/mL, respectively. Throughout our analysis, we used the conversion factor of 5.82 copies/IU for HBV DNA. While standardized HBV RNA assays are not currently available, the in-house assays used to quantify HBV RNA in these trials are linear in the range of HBV RNA observed in trial participants (17). As the analysis below relies on the *relative,* rather than absolute, decline in HBV RNA concentrations, it is unaffected by the use of two different HBV RNA assays in the vebicorvir and ABI-H2158 trials. These assays measure HBV pgRNA in serum or plasma, which we call HBV RNA consistent with previous usage (6).

### Mathematical model

We used a previously developed multiscale mathematical model of chronic HBV infection (18). The model extends the standard viral dynamics model that tracks uninfected hepatocytes (*T*) (19), infected hepatocytes (*I*), and virus (*V*) by explicitly modeling the dynamics of HBV RNA (*R*) and ALT (*A*) in the circulation and the intracellular dynamics of both encapsidated HBV pregenominc RNA, pgRNA (*P*), and relaxed circular DNA, rcDNA (*C*), within infected hepatocytes as was done previously (1,19). Fig. 1 illustrates the model (18), and full mathematical details are given in the SI Appendix.

**Fig. 1.**
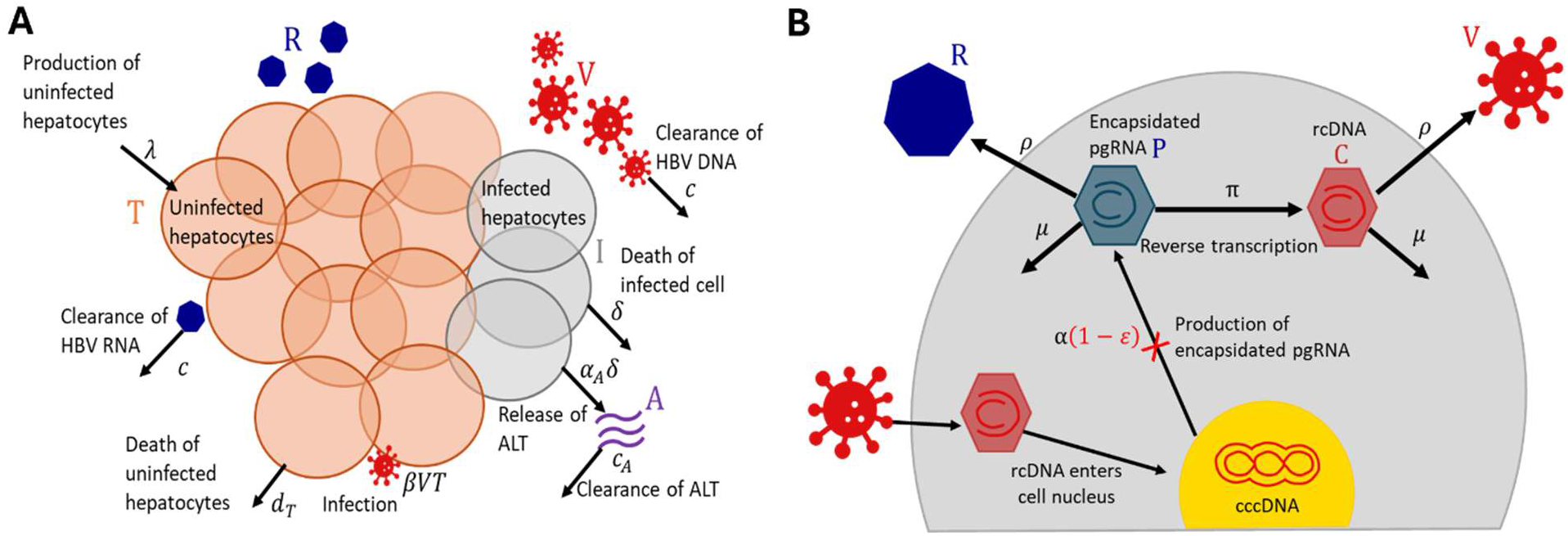
Model schematic showing the extracellular and intracellular mechanisms included in the viral dynamics model. At the extracellular scale, uninfected hepatocytes are produced at a constant rate *λ*, cleared with per capita rate *d*_T_, and infected by HBV at rate *β*. Infected hepatocytes die at per capita rate *δ* and release an amount α_A_ of ALT into the circulation upon death. The background production rate of ALT is given by *s* and circulating ALT is cleared with first order rate constant *c*_A_. Both HBV RNA and HBV DNA are assumed to be cleared from the circulation at per capita rate *c*, as indicated (18). At the intracellular scale, encapsidated pgRNA is produced within infected hepatocytes at rate α, reverse transcribed to form rcDNA with per capita rate *π*, or secreted into the circulation as HBV RNA containing particles at per capita rate *ρ*. Following its synthesis, intracellular rcDNA is assembled into virions and secreted into the circulation also with per capita rate *ρ*. Intracellular encapsidated pgRNA and rcDNA are both assumed to be degraded with per capita rate *μ* (18). This figure is reproduced from Iyaniwura (18) under the Creative Commons Attribution License.

Due to the frequent dosing and rapid pharmacokinetics of these CAMs, we assumed that CAM treatment reduced the production rate of encapsidated pgRNA by a constant factor of (1 − s), where 0 ≤ s ≤ 1 is the CAM effectiveness (18, 20, 21). Here, s = 1 means 100% effectiveness, and s = 0 means no effect. Following cessation of treatment, we used a maximum effect model to capture waning antiviral effects during drug washout (18).

### Data fitting and statistical methods

As the two trials considered different CAMs, treatment durations, and trial populations, we fit the mathematical model to each dataset separately. As done previously, we fixed the model parameters governing uninfected hepatocyte dynamics to literature values (18, 22). For each dataset, we estimated the remaining model parameters using a non-linear mixed-effects approach implemented in Monolix (23) to the totality of circulating HBV RNA, HBV DNA, and ALT data simultaneously. When fitting the model to the vebicorvir and ABI-H2158 trials, we fit 741 and 624 data points, respectively. We previously showed that the model parameters, including the parameters describing intracellular dynamics, are directly informed by this data (18). We left-censored measurements below the LLoD for the vebicorvir trial and the LLoQ for the ABI-H2158 trial.

For both trials, we included dose as a categorical covariate for CAM effectiveness, s. When modeling the vebicorvir trial that included both HBeAg-positive and HBeAg-negative participants, we included HBeAg status as a covariate on the infection rate, *β*, the death rate of infected hepatocytes, *δ*, and the production rate of encapsidated pgRNA, α, as identified in our previous analysis (18). We provide full details of our statistical, covariate, and error models in the SI Appendix.

We used the Mann-Whitney U test to identify any significant differences in the distribution of individual parameter estimates between the two trials. Specifically, for all model parameters except the CAM effectiveness, s, we tested the null hypothesis that the individual parameters obtained by fitting the viral dynamics model to the HBeAg-positive participant data in each trial were drawn from the same distribution. For the CAM effectiveness, s, we tested whether CAM ABI-H2158 was more effective than vebicorvir at a given dose level, as indicated by preclinical experiments (24), and thus used a one-sided test for differences in s between vebicorvir and ABI-H2158 at 100 mg QD and 300 mg QD doses.

## Results

### Model fits to participant data

Our mathematical model captured the dynamics of HBV RNA, HBV DNA, and ALT during treatment and during the viral rebound that followed treatment cessation in both trials. We show the fit of our model to HBV RNA and DNA data from a randomly selected subset of participants from the vebicorvir and ABI-H2158 trials in Figs. 2 and 3, respectively. The model fits to the HBV RNA, HBV DNA, and ALT data for all participants are shown in Figs. S1-S8. We provide population estimates for the CAM effectiveness in Table 1, and the population estimates for all model parameters in Table S1.

**Fig. 2.**
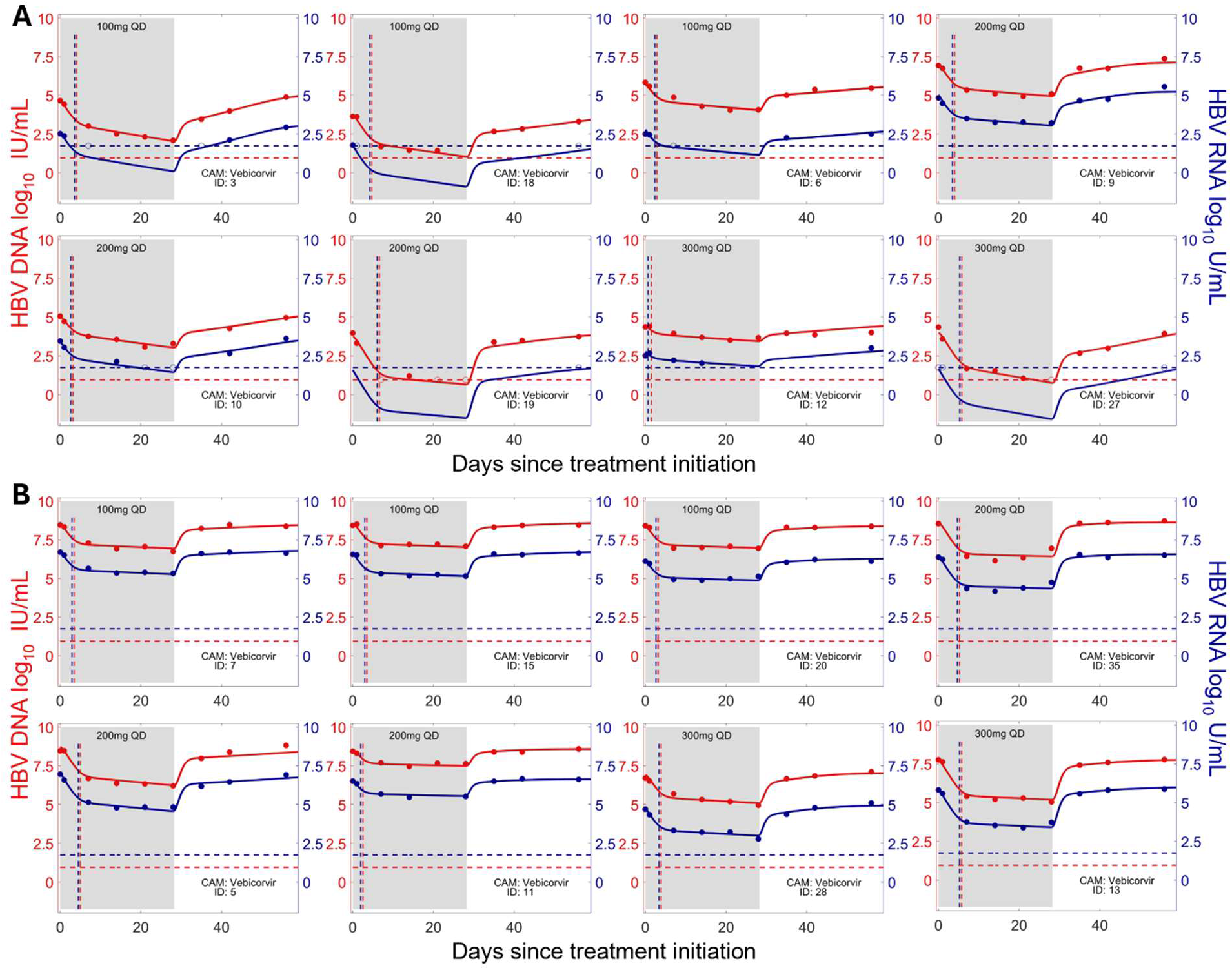
Model fits to HBV RNA (blue) and HBV DNA (red) data from participants in the vebicorvir trial. Model fits for a subset of HBeAg-negative participants (A) and HBeAg-positive participants (B) in the vebicorvir trial. In both cases, the HBV DNA and HBV RNA data are shown as red and blue dots, respectively. The model fits are shown as continuous curves. For each participant, the predicted transition time between the first and second phases of HBV RNA and HBV DNA decline, 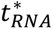 and 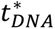, are shown as vertical dashed lines in blue and red, respectively. Model fits to viral load, HBV RNA and ALT for all participants are shown in Figs. S1-S4. For some HBeAg-negative participants, HBV RNA concentrations were below the LLoD (dashed blue line) but were still predicted as the population fitting approach uses data from all participants. The red dashed line is the LLoD for HBV DNA.

**Fig. 3.**
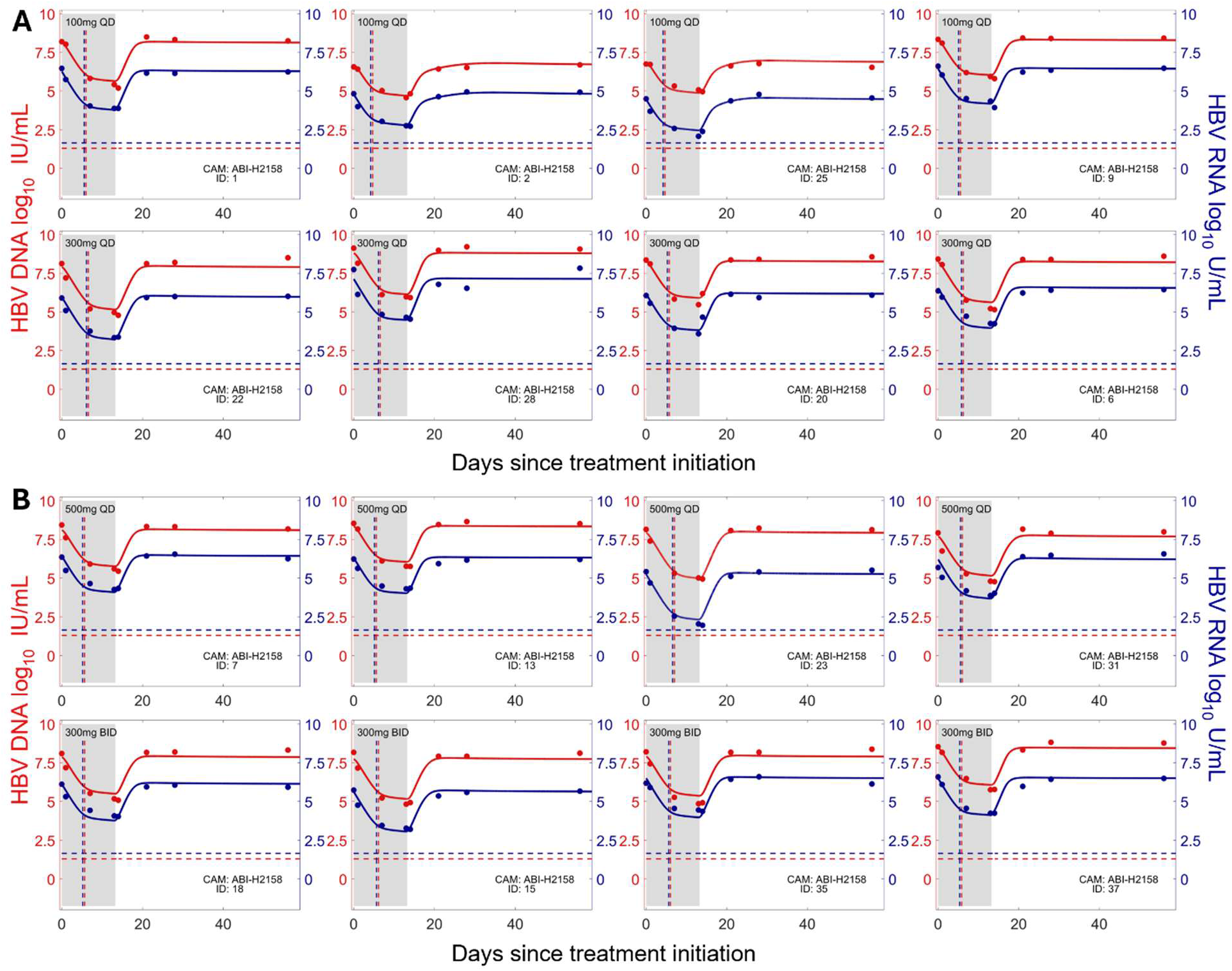
Model fits to HBV RNA and HBV DNA data from participants in the ABI-H2158 trial. (A) The model fits for a subset of participants in the 100 mg QD and 300 mg QD arms of the ABI-H2158 trial. (B) The model fits for a subset of participants in the 500 mg QD and 300 mg BID arms of the ABI-H2158 trial. In both cases, the HBV DNA and HBV RNA data are shown as red and blue dots, respectively, while the corresponding LLoQ is shown as dashed lines. All participants were HBeAg-positive. The model fits are shown as continuous curves. For each participant, the predicted transition time between the first and second phases of HBV RNA and HBV DNA decline, 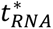 and 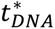, are shown as vertical dashed lines in blue and red, respectively. Model fits to viral load, HBV RNA and ALT for all participants are shown in Figs. S5-S8.

**Table 1.** Estimated CAM effectiveness from fitting the model to the vebicorvir and ABI-H2158 clinical data. NA indicates Not Applicable, because the dose was not administered in the trial.

| Dose level | Vebicorvir effectiveness, $\varepsilon$ | ABI-H2158 effectiveness, $\varepsilon$ |
| --- | --- | --- |
| 100 mg QD | 0.907 | 0.987 |
| 200 mg QD | 0.970 | NA |
| 300 mg QD | 0.984 | 0.995 |
| 500 mg QD | NA | 0.994 |
| 300 mg BID | NA | 0.993 |

We estimated that vebicorvir had a dose-dependent antiviral effect, blocking 90.7%, 97.0%, and 98.4% of encapsidated pgRNA production for the 100 mg, 200 mg, and 300 mg QD dose levels, respectively. Consistent with preclinical characterizations, ABI-H2158 was a more potent inhibitor of encapsidated pgRNA production and was estimated to block 98.7%, 99.5%, 99.4%, and 99.3% of the production for the 100 mg, 300 mg, 500 mg QD and 300 mg BID dose levels, respectively. The similar estimates of s, and thus antiviral effect, for the 300 mg QD, 500 mg QD, and 300 mg BID dose levels of ABI-H2158 are consistent with the less-than-dose-proportional accumulation of this compound (7) and no dose-dependent reduction in log_10_ concentrations of HBV RNA and DNA (7). Altogether, our results indicated that ABI-H2158 was significantly more effective than vebicorvir at both the 100 mg and 300 mg QD doses (*p*-values= 2×10^-4^ and 0.05, respectively, shown in Fig. S9).

Simultaneously fitting both the viral load and ALT dynamics facilitated the estimation of the death rate of infected hepatocytes, *δ* (25). For the HBeAg-positive participants in the vebicorvir trial, we estimated an infected cell half-life of 28.3 days, which is similar to that reported in other studies (26). However, we estimated a significantly shorter half-life of infected cells for participants in the ABI-H2158 trial (half-life of 11.8 days, *p*-value=3×10^-8^, shown in Fig S9). The increased death rate of infected hepatocytes during the ABI-H2158 trial may indicate treatment-induced hepatotoxicity and account for the elevated ALT concentrations that led to the discontinuation of the subsequent phase 2 trial of the ABI-H2158 combination treatment with a NA (7, 27). It is noteworthy that, while the onset of elevated ALT concentrations was generally observed following 2 to 8 weeks of combination treatment (7), our modeling was able to identify this elevated death rate of infected hepatocytes earlier within 14 days of starting ABI-H2158 monotherapy. This is possible because the rates of decline of HBV DNA and HBV RNA during the second phase of viral decline (Figs. 2 and 3) reflect the death rate of infected cells (18).

As shown in Figs. 2 and 3, there was a biphasic decline of both HBV RNA and HBV DNA levels during CAM treatment. Using our model, we calculated the times of transition between the final two phases of HBV RNA and HBV DNA decline during CAM monotherapy, denoted as 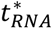 and 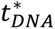 respectively, and detail these calculations in SI Appendix. The model predicted times, 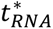and 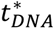, are shown as vertical dashed lines in Figs. 2 and 3. For all participants, 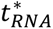 and 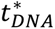 occurred prior to day 7 with 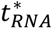 occurring before 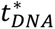 (see SI Appendix for details). This ordering of 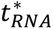 and 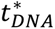 reflects the CAM’s primary mechanism of action of blocking the production of encapsidated pgRNA, as pgRNA that was formed prior to treatment initiation can be reverse transcribed into rcDNA. This rcDNA can then be secreted into the circulation as HBV DNA, which delays the decline of HBV DNA.

### HBV RNA and HBV DNA kinetics directly inform CAM effectiveness

Analogous to a critical observation in the case of direct-acting antivirals against HCV infection (11, 12, 28), we next sought to determine if the decay of HBV RNA and HBV DNA during the first phase of decline could inform CAM effectiveness directly. By analyzing the mathematical model, we show in the SI Appendix that

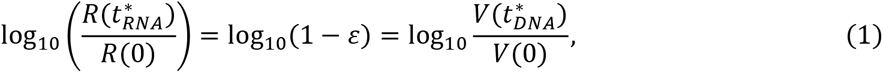

where *R*(0) and *V*(0) are the baseline HBV RNA and HBV DNA concentrations, respectively, 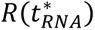 and 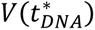 are the HBV RNA and HBV DNA concentrations at the transition from the first to the second phase of declines, and s is the effectiveness of CAM.

We sought to validate this model derived predictive relationship using the viral load data from the vebicorvir and ABI-H2158 trials. The predictive relationship in Eq. (1) links the dynamics of the circulating biomarkers HBV RNA and HBV DNA at the transition between the 1^st^ and 2^nd^ phases of decline with the CAM effectiveness. However, we only have observations to calculate the relative decline in HBV RNA and HBV DNA when these circulating biomarkers were measured, e.g., at day 7 or day 14, and not at 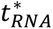 and 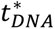 precisely. As mentioned, 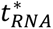 and 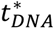 are both near day 7 for participants in the vebicorvir and ABI-H2158 trials. Figure 4A shows a scatter plot of the model estimated values of log_10_ (1-s) versus the relative decline of log_10_ HBV DNA and RNA at day 7 for all participants. The apparent relationship is linear with a slope of 1. We formally tested this prediction by fitting a linear mixed-effects model to the antiviral effectiveness as a function of the relative change in both HBV RNA and HBV DNA from baseline to day 7 for each participant. We used a mixed effects model, rather than simple correlation analysis, as the longitudinal HBV DNA and HBV RNA measurements are not independent (23). The model included a fixed effect for the slope of log-decline and random effects for the intercepts. Eq. (1) predicts that the slope of this model should be unity, and indeed, the estimated slope was 0.99 (95% CI: 0.95–1.02), and not significantly different from one (*p*-value=0.12). This validates Eq. (1) as a predictive relationship between early viral decline and CAM effectiveness. We calculated the coefficient of determination for this linear mixed-effects model (29) and identified a strong (R^2^= 0.80) relationship between the relative HBV RNA and HBV DNA log-decline and CAM effectiveness, expressed as log_10_(1−s). We show the equivalent of Fig. 4A for the individual trials in Fig S10. In all cases, the slope of the linear mixed-effects model is 0.99 and the 95% CI includes unity. We also considered a linear mixed-effects model where the slope was modulated by the biomarker considered, i.e., HBV RNA or HBV DNA. Again, the 95% confidence interval of the grand mean slope included 1 (Fig. S11). The predicted relationship between biomarkers and CAM effectiveness also holds for HBV RNA and HBV DNA, individually (Figs. S12-S13).

**Fig. 4.**
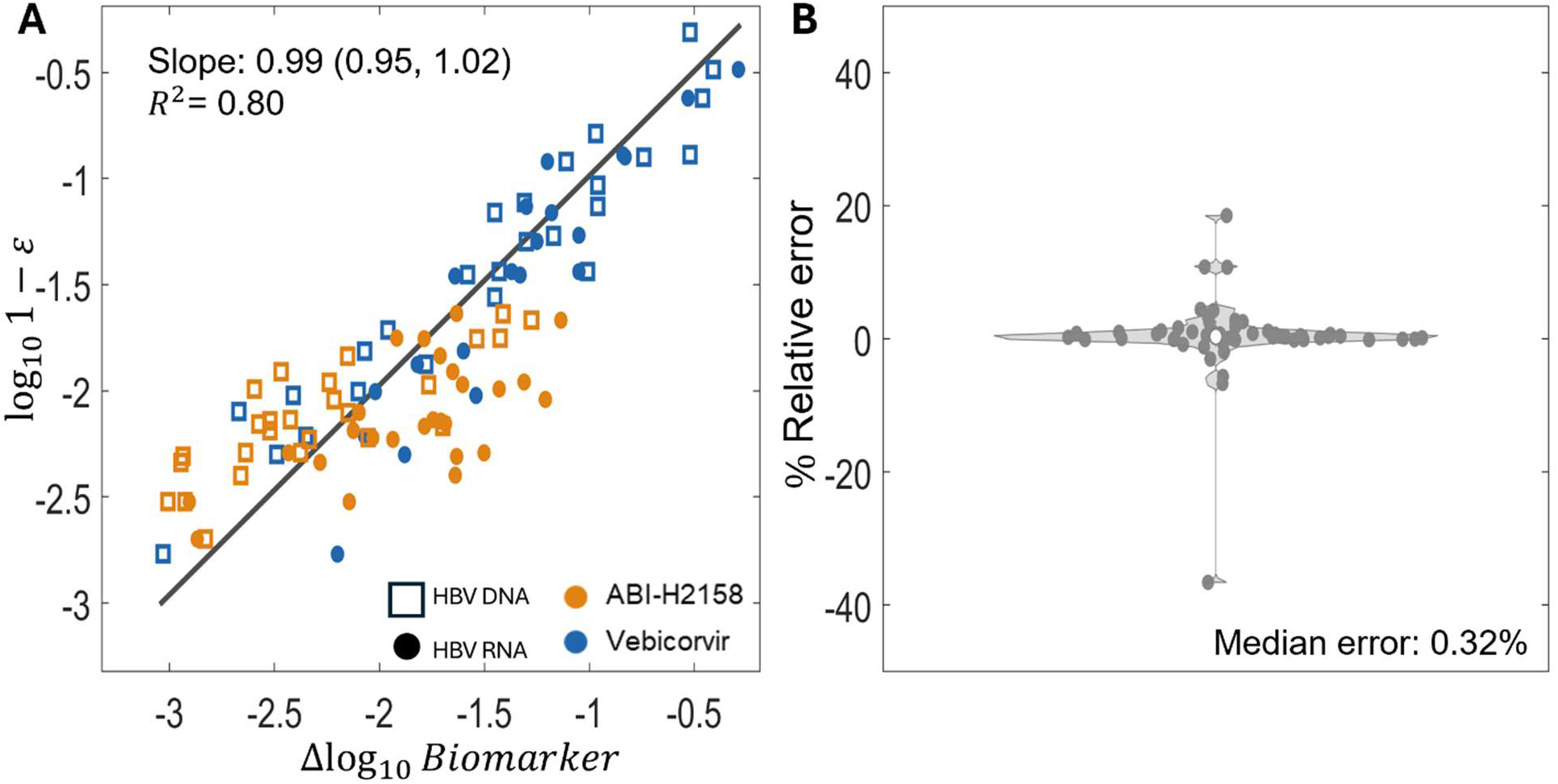
The log-decline of HBV RNA and HBV DNA directly informs CAM effectiveness. (A) A scatter plot of the best-fit individual values of CAM effectiveness, s, expressed as *log*(1 − s), against the observed relative log-decline in HBV RNA and HBV DNA at day 7 in the vebicorvir and ABI-H2158 trials. The linear mixed-effects model has fixed effect (slope) 0.SS with S5% confidence interval and coefficient of determination given in the legend. (B) A violin plot of the percent relative error between the best-fit individual values of CAM effectiveness, s, and the predicted value of s from the relative decay of HBV RNA and HBV DNA for each individual participant in the vebicorvir and ABI-H2158 trials. The median relative error is 0.32%.

Rearranging Eq. (1) gives a simple relationship between s and these circulating biomarkers

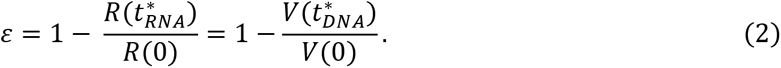

To further test the modeling predictions, we used Eq. (2) to predict the antiviral effectiveness s from the relative decline between baseline and day 7 of HBV RNA and HBV DNA for each participant in the two trials. For each participant, these two biomarkers yield two predictions of s. To test the accuracy of these predictions, we calculated for each individual the percent relative error between the mean of these two predictions and the best-fit value of s, obtained when we fit the full model to the data. The relative decay of HBV RNA and HBV DNA between baseline and day 7 predicted the best-fit CAM effectiveness s, with a median relative error of 0.32%. We show the violin plot of the percent relative error in Fig. 4B, and the corresponding relative error for each individual trial and individual biomarkers in Figs. S14-S16. Together, these results clearly indicate that our proposed method to estimate CAM effectiveness from the decline of HBV RNA and HBV DNA can be useful clinically to quantify the participant-specific CAM effect in inhibiting encapsidated pgRNA production. In fact, our method leads to a simple rule: if there is a 1 log_10_ decline in HBV RNA during the first phase of decay, that is, if *R*(*t*^∗^)/*R*(0) = 10^−1^ then the CAM effectiveness satisfies s = 1 − 0.1 = 0.9. Similarly, a 2 log_10_ decline corresponds to an effectiveness of s = 1 – 0.01 =0.99, etc. The HBV DNA declines obey the same rule.

### HBV RNA and HBV DNA decline during 14 days of monotherapy can be used to evaluate later generation CAMs

An important question during clinical trials is whether a candidate drug is superior to existing drugs. We next tested if our method is sufficiently sensitive to identify this increased effectiveness. We expect later generations of CAMs to be more effective than ABI-H2158, and so we considered a hypothetical next-generation CAM that is 50-fold more effective than 300 mg QD of ABI-H2158, with a population effectiveness estimate of s̅ = 0.9999 = 1 − 10^−4^. We note that the second-generation CAM, ABI-4334, is 500-fold more effective than vebicorvir, as measured in *in vitro* dose-response assays (30), which implies that the high effectiveness of our hypothetical CAM may be observed with next-generation CAMs currently undergoing clinical trials (31). We tested whether the relative decline in HBV RNA and HBV DNA would differentiate this hypothetical CAM from 300 mg QD of ABI-H2158 using virtual clinical trials. We simulated 10,000 trials of 10 virtual participants in each treatment arm to empirically determine the 95% confidence intervals for the estimate of the antiviral effectiveness (see SI Appendix for details). Our method for calculating CAM effectiveness is only approximate if clinical measurements are not made precisely at the transition between the first and second phases of decline. However, day 14 of monotherapy is sufficiently late to capture the entirety of the first phase of HBV RNA and HBV DNA decline, even for the very potent hypothetical CAM, where 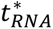 and 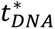 are predicted to be 11 and 12 days, respectively. The additional decline in HBV RNA and HBV DNA between 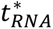 and 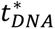 to day 14 may result in slight overestimates of the true antiviral effectiveness. However, our virtual clinical trials indicate that the relative error introduced by this overestimation will be negligible (Figs. S17 and S18).

These simulated clinical trials (Fig. S17) showed that the mean population effectiveness of both ABI-H2158 and the hypothetical CAM are well estimated using Eq. (2). Furthermore, the 95% confidence intervals of the mean population estimates of drug effectiveness calculated using the relative decline in either HBV RNA or HBV DNA clearly differentiate 300 mg QD of ABI-H2158 (CI_95_: [0.993–0.997]) from the hypothetical CAM (CI_95_: [0.99989–0.99995]). Taken together, this indicates that our method can differentiate between highly effective CAMs using only HBV RNA and HBV DNA concentrations collected at baseline and day 14. We also simulated cohorts of sizes varying between N=5 and N=30 and showed that a cohort of 10 participants resulted in enough power to conclude that the hypothetical CAM was significantly more effective than ABI-H2158 (Fig. S18).

Finally, we show, in Fig. S18, that our method for inferring CAM effectiveness from relative decline in HBV RNA and HBV DNA holds true even in the case where the death rate of infected hepatocytes is increased by CAM treatment, as has been suggested for some class A CAMs (4, 32).

## Discussion

HBV viral kinetics during CAM monotherapy is characterized by a biphasic decline in both HBV RNA and HBV DNA (18, 22, 33). In this study, we demonstrated that the magnitude of the decline from baseline to the transition between the two phases of decline of these circulating biomarkers provides a good estimator of CAM effectiveness. We identified a clear predictive relationship between the early dynamics of HBV RNA and HBV DNA and the CAM antiviral effectiveness. This relationship provides a straightforward approach to estimate the *in vivo* effectiveness of next-generation CAMs using short-term clinical trials of approximately two weeks in duration.

Our work supports the role of both HBV RNA and HBV DNA as direct indicators of CAM effectiveness using data from clinical trials of two distinct CAMs. Of these two biomarkers, HBV DNA is traditionally measured in clinical trials of new treatments for chronic HBV infection and thus our results will facilitate evaluation of CAM antiviral effectiveness using standardized and widely accessible assays. While our results immediately extend to estimate the antiviral effectiveness of next-generation CAMs in inhibiting encapsidated pgRNA production tested as monotherapies, our method also applies in the case of CAM and NA combination treatment under the assumption that CAMs and NAs act independently.

It is important to note that both vebicorvir and ABI-H2158 are class-E CAMs that induce the formation of empty capsids rather than the aberrant capsids produced during class-A CAM treatment (34). Both classes of CAMs inhibit the production of encapsidated pgRNA, which then drives the first phase of decline observed during CAM monotherapy. However, in HBV mouse models, some class-A CAMs have been shown to increase the death rate of infected hepatocytes (4, 32), although this secondary antiviral effect may require high levels of intracellular HBcAg (35). While not yet shown in the clinic, if class-A CAMs had this effect in humans, the treatment-mediated death of infected hepatocytes would drive a faster second phase of decline of HBV RNA and HBV DNA and may lead to an overestimation of the CAM effectiveness using our method.

Nevertheless, our clinical trial simulations discussed in the SI Appendix indicate that the relative decline in HBV RNA or HBV DNA between baseline and day 14 would only overestimate the effectiveness of a hypothetical class-A CAM by less than 0.01% (Fig. S18), even in the setting of a treatment mediated higher death rate. Consequently, we expect that the predictive relationship between the relative decline of HBV RNA and HBV DNA with CAM effectiveness will hold, even for next-generation CAM-As.

HBV RNA is a direct downstream product of cccDNA that is unaffected by NA treatment. Consequently, HBV RNA dynamics have been proposed as a marker of cccDNA transcriptional activity during NA treatment (33, 36, 37). However, this is not the case for CAM treatment, as we and others (18, 22) have shown that HBV RNA dynamics during CAM treatment reflect CAM-mediated blocking of the production of encapsidated pgRNA, rather than an inhibition of cccDNA transcriptional activity. Thus, establishing a link between cccDNA activity and other circulating biomarkers such as HBcrAg, HBeAg and HBsAg during early CAM treatment will be necessary (38–40). Identifying an informative biomarker of cccDNA activity during CAM treatment will be particularly important in trials of next-generation CAMs that have demonstrated *in vitro* inhibition of cccDNA formation (30, 41).

Nevertheless, we expect our main result linking early HBV RNA decline and CAM effectiveness to hold for next-generation CAMs that both inhibit production of encapsidated pgRNA and cccDNA formation. Specifically, given the relatively long half-life of cccDNA (42), the first phase of HBV RNA and DNA decline in trials of these next-generation CAMs should be driven by the inhibition of pgRNA production by infected hepatocytes rather than reductions of the intracellular cccDNA pool. However, the second phase of HBV RNA decline, which our results clearly link to the loss of infected hepatocytes (18, 22), would then reflect the loss of infected hepatocytes producing HBV DNA and HBV RNA due both to death of these cells and loss of cccDNA within infected hepatocytes during treatment (43). Therefore, we expect that viral kinetic modeling, such as extensions of the work by Kitagawa et al. (43), will prove useful in quantifying the effect of next-generation CAMs in blocking cccDNA formation. Overall, our results not only permit the straightforward evaluation of the antiviral effectiveness of current CAMs during the first 14 days of treatment but will also facilitate the quantification of this secondary mechanism of action on cccDNA that will be critical for HBV functional cure. Furthermore, attaining a cure or functional cure of HBV will most likely require a combination of antiviral and immunostimulatory agents.

Biological insights obtained from this modeling may facilitate the evaluation of next-generation therapies with multiple mechanisms of action, and our results illustrate how mechanistic modeling can be used to extract quantitative insight from early clinical data.

## Data Availability

All data produced in the present study are available upon reasonable request to the authors

## Acknowledgments

We thank Assembly Biosciences for supplying the data used in this analysis, and Carolin Zitzmann for assistance with the study.

## Notes

### Competing Interest Statement

The authors have declared no competing interest.

### Clinical Trial

NCT02908191, NCT03714152

### Author Declarations

This research was approved by the Los Alamos National Laboratory Human Subjects Research Review Board.

### Funding

- National Institutes of Health: R01-AI078881, R01-OD011095, R01-AI116868
- United States Department of Energy: 89233218CNA000001

