## Supplemental information for "The magnitude of early hepatitis B RNA and DNA declines directly inform capsid assembly modulator effectiveness"

#### Multiscale model of chronic hepatitis B infection

In prior work, we developed a multiscale model of chronic HBV infection [7]. While standard viral dynamics models focus on the dynamics of infected and uninfected hepatocytes and circulating HBV RNA and DNA, we explicitly include the intracellular dynamics of HBV pgRNA and rcDNA through a time-since-infection model. In the model, uninfected hepatocytes, $T$, are produced at a constant rate $\lambda$, cleared at per capita rate $d_T$, and infected at rate $\beta$. These infected hepatocytes are cleared with a per capita death rate $\delta$. As we do not explicitly model the cccDNA dynamics within infected hepatocytes, we model the production of encapisdated pgRNA at a constant rate $\alpha$ under the assumption that the cccDNA concentration within infected cells does not vary during the short duration of these phase I trials. This intracellular encapsidated pgRNA is reversed transcribed to form intracelluar HBV rcDNA at a per capita rate $\pi$. Both encapsidated HBV pgRNA and rcDNA decay, with per capita rate $\mu$, and are exported into the circulation as HBV RNA or HBV DNA containing particles with per capita rates $\rho_r$ and $\rho_v$, respectively, where we impose $\rho_r = \rho_v = \rho$ in our fitting. A model diagram is given in Fig. 1 of the main text. Finally, we use the Cardozo et al. [2] model for the dynamics of ALT, $A(t)$, which was based on the ALT model in Ribeiro et al. [11].

As we show in Iyaniwura et al. [7], if the model parameters are independent of the duration of time that a cell has been infected, the time-since-infection model can be formulated as the system of ordinary differential equations (ODEs). The ODE model describes the dynamics of uninfected and infected target cells, as well as circulating HBV

RNA, HBV DNA, and ALT, and is given by

$$\left.\begin{aligned}
\frac{\mathrm{d}}{\mathrm{dt}}T(t) &= \lambda - \beta VT - d_T T,\\
\frac{\mathrm{d}}{\mathrm{dt}}I(t) &= \beta VT - \delta I,\\
\frac{\mathrm{d}}{\mathrm{dt}}P(t) &= \alpha(1-\varepsilon(t))I - (\mu+\delta+\pi+\rho_r)P,\\
\frac{\mathrm{d}}{\mathrm{dt}}C(t) &= \pi P - (\mu+\delta+\rho_v)C,\\
\frac{\mathrm{d}}{\mathrm{dt}}R(t) &= \rho_r P - c_r R,\\
\frac{\mathrm{d}}{\mathrm{dt}}V(t) &= \rho_v C - c_v V\\
\frac{\mathrm{d}}{\mathrm{dt}}A(t) &= s + \alpha_A \delta I - c_A A.
\end{aligned}\right\} \tag{S1}$$

In Eq. (S1), $T(t)$ is the concentration of uninfected hepatocytes, while $I(t)$ represents the total concentration of infected hepatocytes, while $V(t)$ and $R(t)$ represent the concentration of HBV RNA and HBV DNA measured in the circulation. In the ODE formulation, $P(t)$ and $C(t)$ represent the total amount of encapsidated pgRNA and rcDNA within infected hepatocytes, respectively.

#### Explicit expression for the transition between first and second phase of decay

We analyze the viral dynamics model Eq. (S1) to characterize the dynamics of HBV RNA during CAM monotherapy. Our analysis relies on the common assumption that new infections are negligible under highly effective antiviral treatment [5, 6]. We previously showed that vebicorvir, and thus ABI-H2158, is sufficiently potent to neglect new infections during the first 10 days of treatment [7]. Under this assumption, the HBV RNA and HBV DNA dynamics are given by a sum of exponential terms. We now analyze the explicit expression for $R(t)$ to derive an explicit expression for the time of the transition between the first and second phase of HBV RNA decline, $t^*_{RNA}$.

To simplify the notation in the upcoming analysis, we denote the loss rate of intracellular pgRNA by

$$\xi_p = \mu + \delta + \pi + \rho.$$

Then, as shown in Iyaniwura et al. [7], the total amount of intracellular pgRNA satisfies

$$P(t) = P_0 e^{-\xi_p t} + \frac{\alpha(1-\varepsilon)I_0}{\xi_p - \delta}\left(e^{-\delta t} - e^{-\xi_p t}\right).$$

where $P_0$ and $I_0$ are the baseline levels of encapsidated pgRNA within infected cells and concentration of infected cells, respectively, corresponding to the steady-state pre-treatment initial conditions. Accordingly, the dynamics of circulating HBV RNA are given by [7]

$$R(t) = Ae^{-ct} + Be^{-\xi_p t} + Ce^{-\delta t}. \tag{S2}$$

As the system was in steady-state prior to treatment, these constants are given by

$$A = R_0 - B - C, \quad B = \frac{cR_0}{c-\xi_p}\left(1 - \frac{(1-\varepsilon)\xi_p}{\xi_p - \delta}\right), \quad \text{and} \quad C = \left(\frac{cR_0}{c-\delta}\right)\left(\frac{(1-\varepsilon)\xi_p}{\xi_p-\delta}\right). \tag{S3}$$

Thus, on the log scale, we have

$$\log(R(t)) = \log\left(Ae^{-ct} + Be^{-\xi_p t} + Ce^{-\delta t}\right).$$

While it appears that HBV RNA should exhibit tri-phasic decay on the log scale, the parameter estimates in Table S1 and identifiability analysis in Iyaniwura et al. [7] imply that

$$\xi_p > \pi = \frac{V(0)(\rho+\delta)}{R(0)} \approx 200 \text{ d}^{-1}.$$

Consequently, the term $B\exp(-\xi_p t)$ decays to zero extremely rapidly, so only two phases of HBV RNA decline are observed during CAM treatment. Thus, following the unobservable initial phase of decline at rate $\xi_p$, a good approximation of the dynamics of HBV RNA is

$$\log(R(t)) = \log(Ae^{-ct} + Ce^{-\delta t}). \tag{S4}$$

where $A > 0$ and $C > 0$ are given in Eq. (S3).

We now can derive an explicit expression for the transition between the first and second phases of decline. From Eq. (S4), these phases of decline have approximate slopes $-c$ and $-\delta$, respectively. The transition time, $t^*_{RNA}$, between the first and second phase of decline can be obtained by equating $Ae^{-ct^*_{RNA}} = Ce^{-\delta t^*_{RNA}}$, i.e. the point at which the extrapolated first phase decline intersects the extrapolated 2nd phase decline. We find

$$t^*_{RNA} = \frac{\log\left(\frac{A}{C}\right)}{c-\delta}. \tag{S5}$$

Iyaniwura et al. [7] showed that the HBV DNA dynamics are given by a sum of four exponentials that reflect the decay of intracellular pgRNA and rcDNA concentrations, with rates $\xi_p$ and $\xi_c = \mu + \delta + \rho$, and the natural decay rate of circulating HBV DNA, $c$, and the death of infected cells, $\delta$. Once again, the decay of intracellular pgRNA is too fast to observe, so HBV DNA dynamics satisfy

$$V(t) = (V_0 - D - E)\, e^{-ct} + De^{-\xi_c t} + Ee^{-\delta t},$$

where

$$D = \frac{\rho}{(c-\xi_c)(\xi_c-\delta)}\left(C_0(\xi_c-\delta) - \frac{\alpha(1-\varepsilon)\pi(1-\varepsilon_{NA})I_0}{\xi_p-\delta}\right) \quad \text{and} \quad E = \frac{\rho\alpha(1-\varepsilon)\pi(1-\varepsilon_{NA})I_0}{(\xi_p-\delta)(c-\delta)(\xi_c-\delta)}.$$

The decay of intracellular rcDNA leads to a transient shoulder in HBV DNA concentrations following treatment initiation before the beginning of biphasic declines. We proceed analogously to our derivation of $t^*_{RNA}$ by equating

$$(V_0 - D - E)\, e^{-ct^*_{DNA}} = Ee^{-\delta t^*_{DNA}}$$

to find

$$t^*_{DNA} = \frac{\log(V_0 - D - E) - \log(E)}{c-\delta}. \tag{S6}$$

In forthcoming work, we use a perturbation argument to show that the transient shoulder in HBV DNA dynamics has negligible impact on the value of $t^*_{DNA}$.

#### Derivation of Eq. (1)

We now derive the relationship between log decline in HBV RNA and $\varepsilon$ given in Eq. (1) of the Main Text. The analysis for HBV DNA is similar and uses the explicit expression for $t^*_{DNA}$.

During therapy, the HBV RNA dynamics on the $\log_{10}$ scale are given, to a good approximation, by

$$\log_{10}(R(t)) = \log_{10}(Ae^{-ct} + Ce^{-\delta t}).$$

Using the definition of $t^*_{RNA}$, we must have $Ae^{-ct^*_{RNA}} = Ce^{-\delta t^*_{RNA}}$ and so

$$\log_{10}(R(t^*_{RNA})) = \log_{10}\left[2C\left(\frac{A}{C}\right)^{\frac{-\delta}{c-\delta}}\right],$$

where we have substituted Eq. (S5) for $t^*_{RNA}$. Simplifying yields the explicit expression for the HBV RNA concentration at the transition between the first and second phase of decline

$$\log_{10}(R(t^*_{RNA})) = \log_{10}\left(2A^{-\frac{\delta}{c-\delta}}\right) + \frac{c}{c-\delta}\log_{10}(C). \tag{S7}$$

The parameter values given in Table S1 satisfy $\delta \ll c \ll \xi_p$. Therefore, in the following analysis, we approximate

$$\frac{c}{c-\delta} \approx 1, \quad \frac{\xi_p}{\xi_p - \delta} \approx 1, \quad \text{and} \quad \frac{\xi_p}{\xi_p - c} \approx 1, \tag{S8}$$

which gives

$$A \approx R_0 \left( 1 - \frac{c}{c - \xi_p} \varepsilon - (1 - \varepsilon) \right) = R_0 \left( \varepsilon \left( \frac{\xi_p}{\xi_p - c} \right) \right).$$

For highly effective CAMs, $\varepsilon \approx 1$, so $A \approx R_0$. The first term in Eq. (S7) then becomes

$$\log_{10} \left[ 2 A^{-\frac{\delta}{c-\delta}} \right] = -\frac{\delta}{c - \delta} \log_{10} \left[ 2^{\frac{\delta - c}{\delta}} R_0 \right].$$

We now estimate this term using the population parameter values for the ABI-H2158 and vebicorvir trials. In all cases, $c = 1$. The mean pre-treatment HBV RNA concentration in the ABI-H2158 trial was $R(0) = 10^{6.7} \approx A$ copies/mL and $\delta \approx 0.05 = 1/20$/day. In the vebicorvir trial, the mean pre-treatment HBV RNA concentration was $R(0) = 10^{6.9}$ copies/mL among HBeAg-positive participants and $\delta = 0.025 = 1/40$/day. For HBeAg-negative participants in the vebicorvir trial, the mean baseline HBV RNA concentration was $R(0) = 10^{3.7}$ copies/mL and $\delta = 0.07 = 7/100$/day in participants with baseline HBV RNA above the LLoQ. Then, for the ABI-H2158 trial and HBeAg-positive and HBeAg-negative participants in the vebicorvir trials, respectively, we estimate

$$\begin{cases} \frac{\delta}{c-\delta} \log_{10} \left[ 2^{\frac{\delta - c}{\delta}} A \right] \approx \frac{1}{19} \log_{10} \left[ 2^{-19} 10^{6.7} \right] \approx 5 \times 10^{-2}, \\ \frac{\delta}{c-\delta} \log_{10} \left[ 2^{\frac{\delta - c}{\delta}} A \right] \approx \frac{1}{39} \log_{10} \left[ 2^{-39} 10^{6.9} \right] \approx -1 \times 10^{-1}, \\ \frac{\delta}{c-\delta} \log_{10} \left[ 2^{\frac{\delta - c}{\delta}} A \right] \approx \frac{7}{93} \log_{10} \left[ 2^{-93/7} 10^{3.7} \right] \approx -2 \times 10^{-2}. \end{cases}$$

In all cases, these are negligible compared to the second term in Eq. (S7). Then, the expression for $C$ immediately yields

$$\log_{10}(R(t^*)) = \log_{10}(R_0) + \log_{10}(1 - \varepsilon),$$

and rearranging gives Eq. (1) of the main text

$$\log_{10}(R(t^*)/R_0) = \log_{10}(1 - \varepsilon). \tag{S9}$$

#### CAM pharmacokinetics

In the studies we consider, CAMs are administered at least daily during the treatment period $[0, \tau]$ and rapidly reach steady-state concentrations [1, 15]. Rather than using a detailed pharmacokinetic model to drive CAM mediated inhibition of encapisdated pgRNA production, we decrease $\alpha$ by a constant factor of $(1-\varepsilon)$ during daily treatment. This approach is common in viral dynamics models [3, 4, 6, 12]. Following the cessation of treatment on day $t=\tau$, we use a maximal effect model to capture the waning drug effect during drug-washout. Specifically, we assume the CAM has a terminal half-life of $t_{1/2}=\log_e(2)/k$, where we estimate the rate $k$. Then, by enforcing continuity in the CAM effect at time $t=\tau$, the effects of CAM treatment are given by [7]

$$\varepsilon(t)=\begin{cases}\varepsilon & \text{if} \quad t\leq\tau,\\ \frac{\varepsilon\exp(-k(t-\tau))}{\varepsilon(\exp(-k(t-\tau))-1)+1}, & \text{if} \quad t>\tau.\end{cases} \tag{S10}$$

### Structural, covariate, and error models for parameter estimation

We estimated model parameters by fitting viral load data from both clinical trials using the non-linear mixed effects modeling framework implemented in Monolix [8, 13]. The details of the structural and error models used in our fitting are given in Iyaniwura et al. [7]. The vebicorvir trial included both HBeAg-positive and HBeAg-negative participants. To account for the mechanistic differences between HBeAg-positive and HBeAg-negative infection, we included HBeAg covariates on the model parameters $\beta, \alpha$, and $\delta$ when fitting our model to the vebicorvir trial [7].

#### Population parameters

We estimated the population parameters using a nonlinear mixed-effects modelling framework implemented in Monolix Version 2024. We report the population level parameter estimates here for both trials. In general, all parameters were well-estimated using the nonlinear mixed-effects framework. As we have previously shown, the model parameters are directly informed by the circulating HBV RNA, HBV DNA, and ALT dynamics under the assumption of chronic infection [7].

### Model fits to individual participants

In the main text, we showed the model fits to the viral load data from a subset of participants in both the vebicorvir and ABI-H2158 trials. Here, we show the model fits to both HBV RNA and HBV DNA for all HBeAg-negative and positive participants of the vebicorvir trial in Figures S1 and S2, respectively, with the ALT fits shown for these participants in Figures S3 and S4. The HBV RNA and HBV DNA dynamics for all participants in the ABI-H2158, separated by dose cohort, are shown in Figures S5 and S6. The ALT fits for these participants are shown in Figures S7 and S8.

| **Parameter** (units) | **Vebicorvir** Fixed effect (RSE) | **Vebicorvir** Random effect (RSE) | **ABI-H2158** Fixed effect (RSE) | **ABI-H2158** Random effect (RSE) |
|---|---|---|---|---|
| $\varepsilon$ (100 mg QD) | 0.907 (4.6) | 1.355 (14.3) | 0.987 (0.4) | 0.61 (23.0) |
| $\varepsilon$ (200 mg QD) | 0.970 (1.5) | 1.355 (14.3) | NA | NA |
| $\varepsilon$ (300 mg QD) | 0.984 (0.9) | 1.355 (14.3) | 0.995 (40.4) | 0.61 (23.0) |
| $\varepsilon$ (500 mg QD) | NA | NA | 0.994 (72.6) | 0.61 (23.0) |
| $\varepsilon$ (300 mg BID) | NA | NA | 0.993 (55.4) | 0.61 (23.0) |
| $\beta$ (mL/copies/day) | | | | |
| HBeAg-negative | $4.21 \times 10^{-7}$ (4.5) | 0.92 (15.2) | NA | NA |
| HBeAg-positive | $1.01 \times 10^{-10}$ (2.9) | - | $7.8 \times 10^{-9}$ (3.2) | 0.16 (68.4) |
| $\alpha$ (copies/cell/day) | | | | |
| HBeAg-negative | 0.20 (34.3) | 0.81 (15.7) | NA | NA |
| HBeAg-positive | 390.84 (9.2) | 0.81 (15.7) | 633.87 (4.2) | 0.53 (15.5) |
| $\pi$ (1/day) | 204.64 (5.57) | 0.35 (5.5) | 187.07 (7.5) | 0.38 (15.9) |
| $\delta$ (1/day) | | | | |
| HBeAg-negative | 0.070 (14.4) | 0.38 (34.1) | NA | NA |
| HBeAg-positive | 0.025 (28.9) | 0.38 (34.1) | 0.059 (13.7) | 0.136 (NaN) |
| $\rho$ (1/day) | 2.48 (26.6) | Fixed | 2.61 (36.7) | Fixed |
| $c$ (1/day) | 1 (-) | Fixed | 1 (-) | Fixed |
| $\mu$ (1/day) | 0 (-) | Fixed | 0 (-) | Fixed |
| $c_A$ (1/day) | 0.057 (26.4) | Fixed | 0.28 (91.8) | Fixed |
| $A_0$ (U/L) | 38.37 3.5) | 0.30 (13.6) | 30.48 (3.2) | 0.24 (14.8) |
| $A_{ue}$ (U/L) | 22.80 (5.5) | 0.27 (20.1) | 19.63 (18.1) | 0.45 (37.5) |
| $k$ (1/day) | 2.29 (230) | 0.24 (71.6) | 1.5 (9.3) | 0.058 (112) |

Table S1: **Population parameters estimated by fitting Eq.** (S1) **to the vebicorvir and ABI-H2158 viral load data.** RSE is the percent relative standard error in the estimate of each parameter. NA indicates that the parameter was not estimated for this trial, while parameters that were estimated at the population level without inter-individual variability are denoted as fixed. In the Vebicorvir trial, we identified a significant negative correlation between $\beta$ and $\alpha$ with correlation coefficient of $-0.95$.

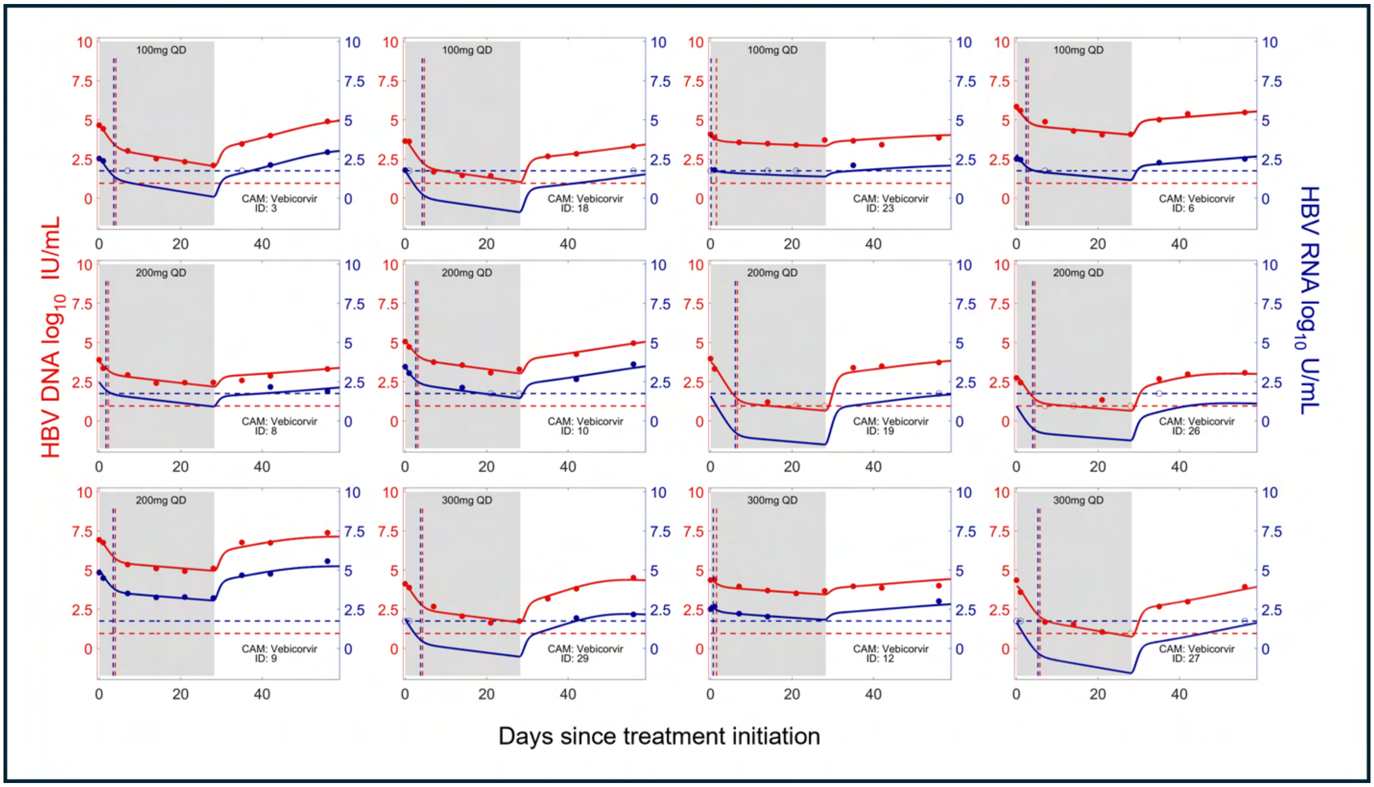


Figure S1: **The model fits to viral load data for HBeAg-negative participants in the vebicorvir trial.** The HBV RNA and HBV DNA data are shown as blue and red circles, respectively. The model fits are shown as continuous curves and the blue and red dashed lines represent the lower limit of detection of the HBV RNA and HBV DNA assays, respectively. For each participant, the predicted transition time between the first and second phase of decline of HBV RNA, $t^*_{RNA}$, and HBV DNA, $t^*_{DNA}$, are shown as vertical dashed lines in blue and red, respectively.

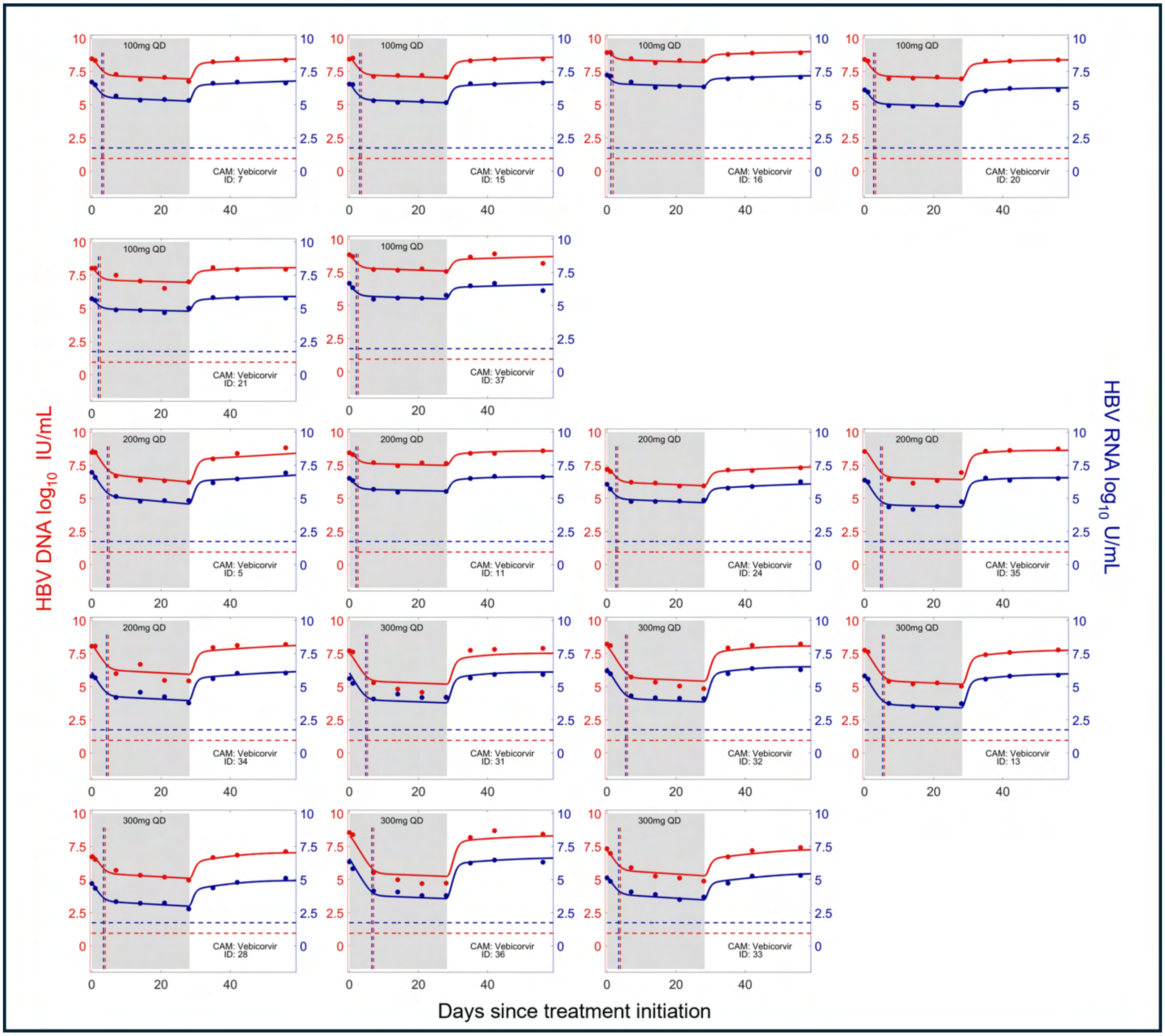


Figure S2: **The model fits to viral load data for HBeAg-positive participants in the vebicorvir trial.** The HBV RNA and HBV DNA data are shown as blue and red circles, respectively. The model fits are shown as continuous curves and the blue and red dashed lines represent the lower limit of detection of the HBV RNA and HBV DNA assays, respectively. For each participant, the predicted transition time between the first and second phase of decline of HBV RNA, $t^*_{RNA}$, and HBV DNA, $t^*_{DNA}$, are shown as vertical dashed lines in blue and red, respectively.

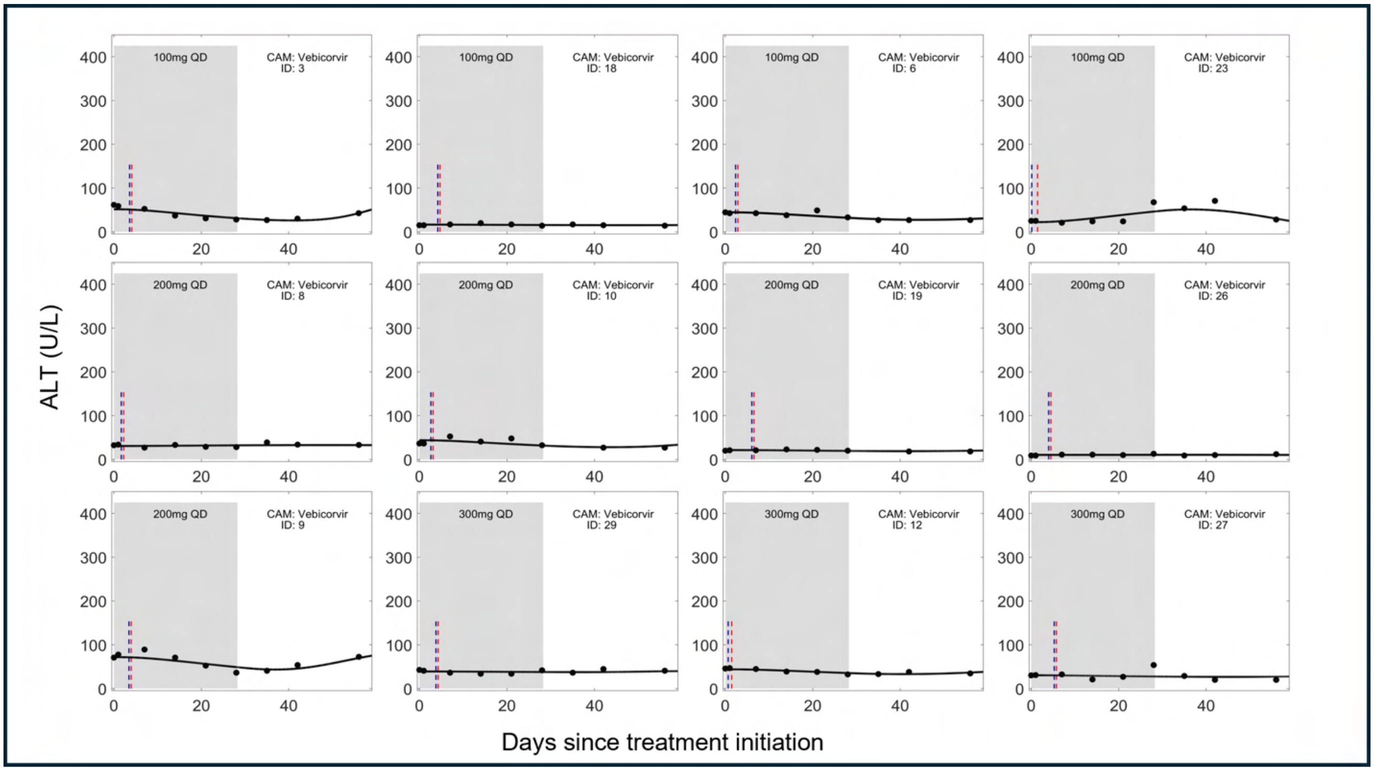


Figure S3: **The model fits to ALT data for HBeAg-negative participants in the vebicorvir trial.** The ALT concentrations are shown as black circles. The model fits are shown as continuous curves. For each participant, the predicted transition time between the first and second phase of decline of HBV RNA, $t^*_{RNA}$, and HBV DNA, $t^*_{DNA}$, are shown as vertical dashed lines in blue and red, respectively.

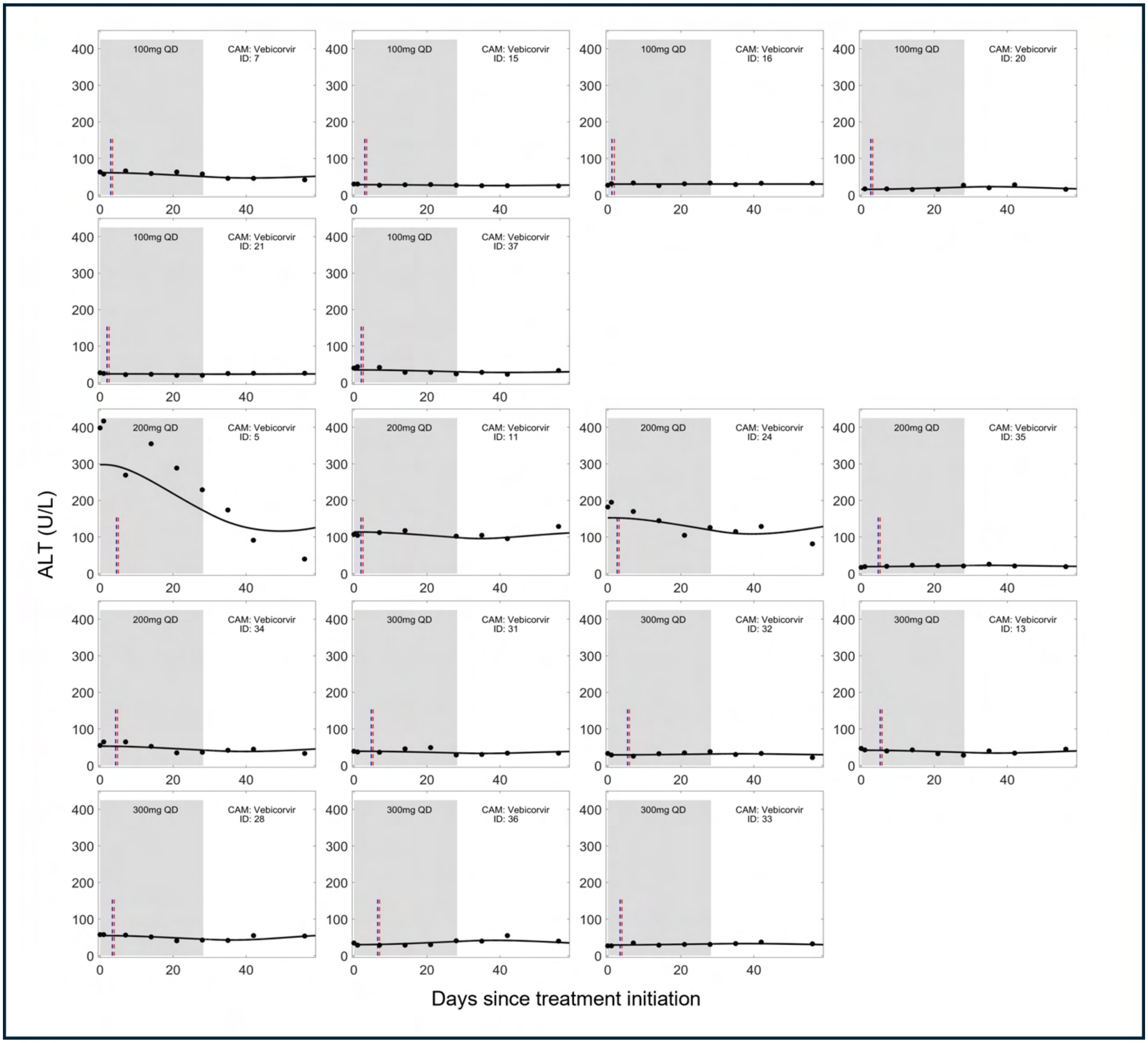


Figure S4: **The model fits to ALT data for HBeAg-positive participants in the vebicorvir trial.** The ALT concentrations are shown as black circles. The model fits are shown as continuous curves. For each participant, the predicted transition time between the first and second phase of decline of HBV RNA, $t^*_{RNA}$, and HBV DNA, $t^*_{DNA}$, are shown as vertical dashed lines in blue and red, respectively.

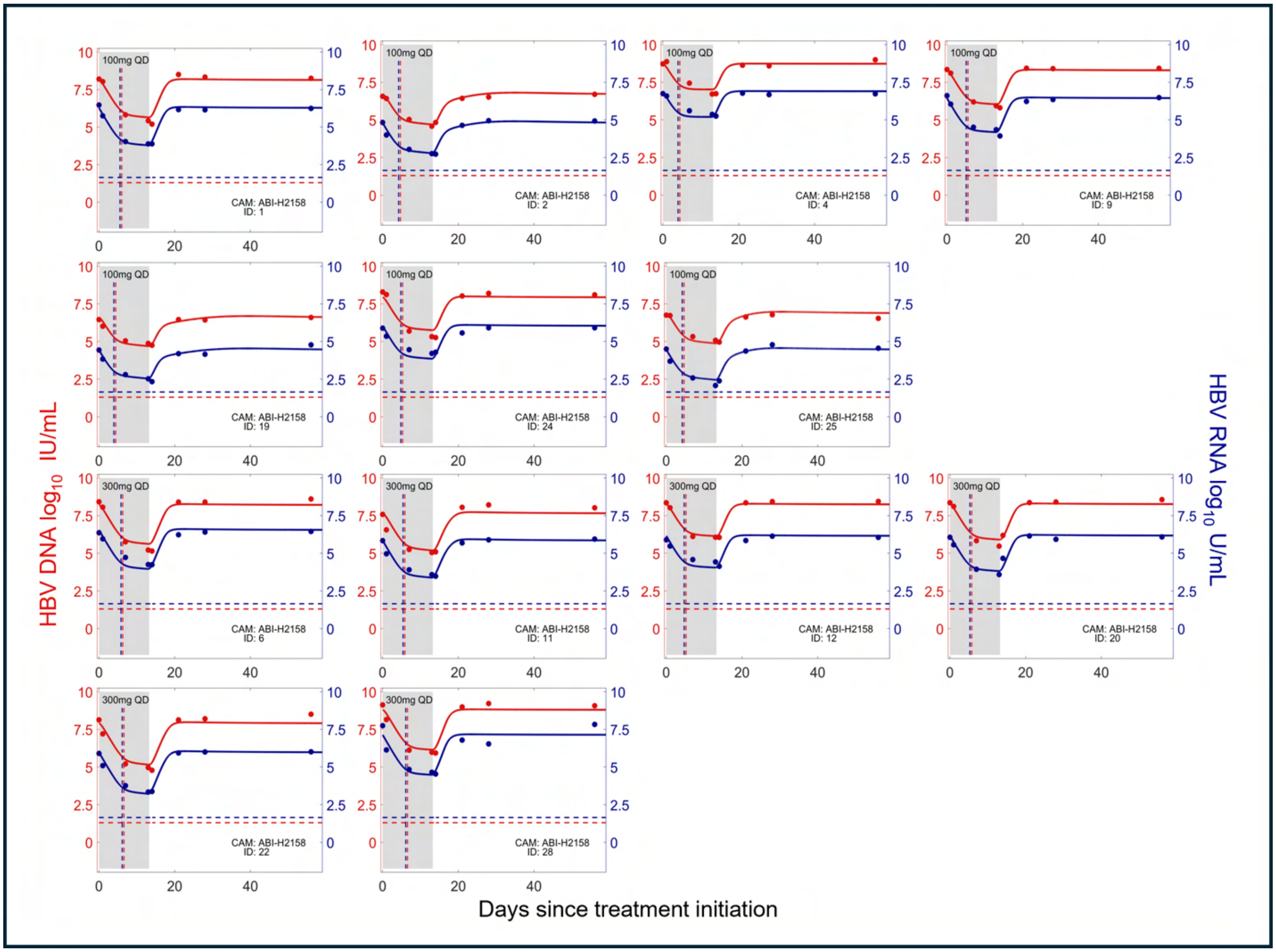


Figure S5: **The model fits to viral load data for participants in the 100 mg QD and 300 mg QD cohorts of the ABI-H2158 trial.** The HBV RNA and HBV DNA data are shown as blue and red circles, respectively. The model fits are shown as continuous curves and the blue and red dashed lines represent the lower limit of quantitation of the HBV RNA and HBV DNA assays, respectively. For each participant, the predicted transition time between the first and second phase of decline of HBV RNA, $t^*_{RNA}$, and HBV DNA, $t^*_{DNA}$, are shown as vertical dashed lines in blue and red, respectively.

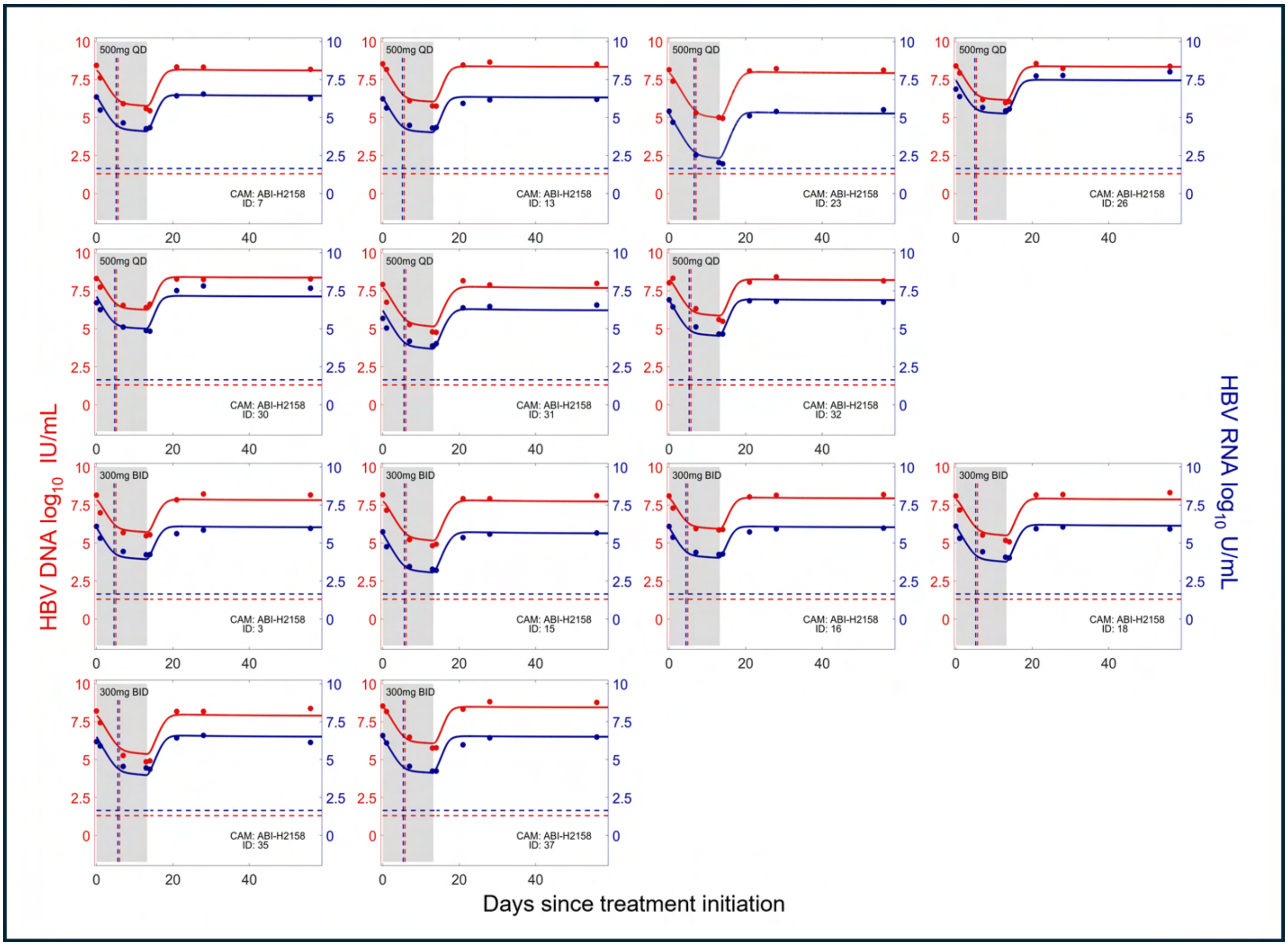


Figure S6: **The model fits to viral load data for participants in the 500 mg QD and 300 mg BID cohorts of the ABI-H2158 trial.** The HBV RNA and HBV DNA data are shown as blue and red circles, respectively. The model fits are shown as continuous curves and the blue and red dashed lines represent the lower limit of quantitation of the HBV RNA and HBV DNA assays, respectively. For each participant, the predicted transition time between the first and second phase of decline of HBV RNA, $t^*_{RNA}$, and HBV DNA, $t^*_{DNA}$, are shown as vertical dashed lines in blue and red, respectively.

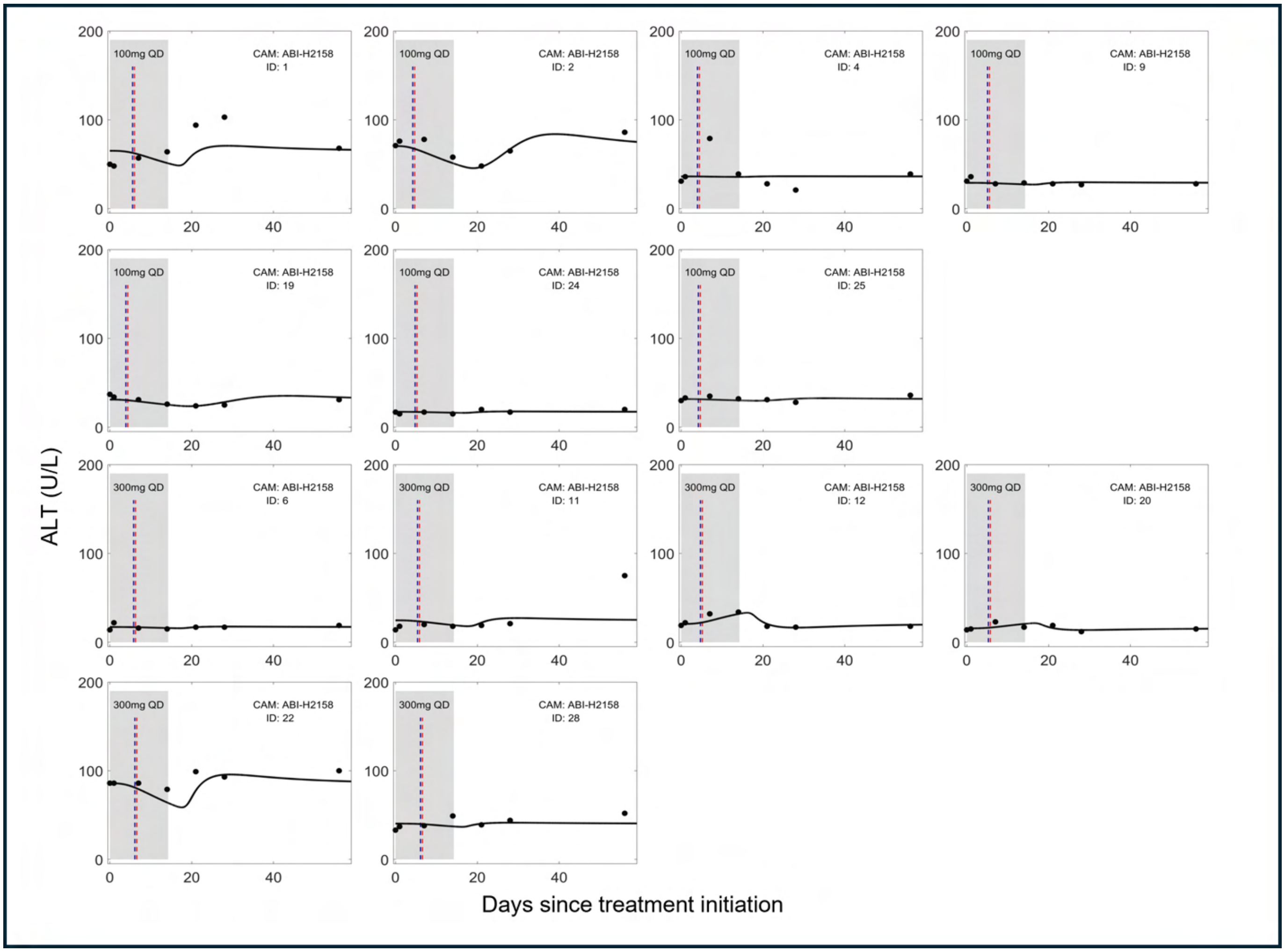


Figure S7: **The model fits to ALT data for participants in the 100 mg QD and 300 mg QD cohorts of the ABI-H2158 trial.** The ALT concentrations are shown as black circles. The model fits are shown as continuous curves. For each participant, the predicted transition time between the first and second phase of decline of HBV RNA, $t^*_{RNA}$, and HBV DNA, $t^*_{DNA}$, are shown as vertical dashed lines in blue and red, respectively.

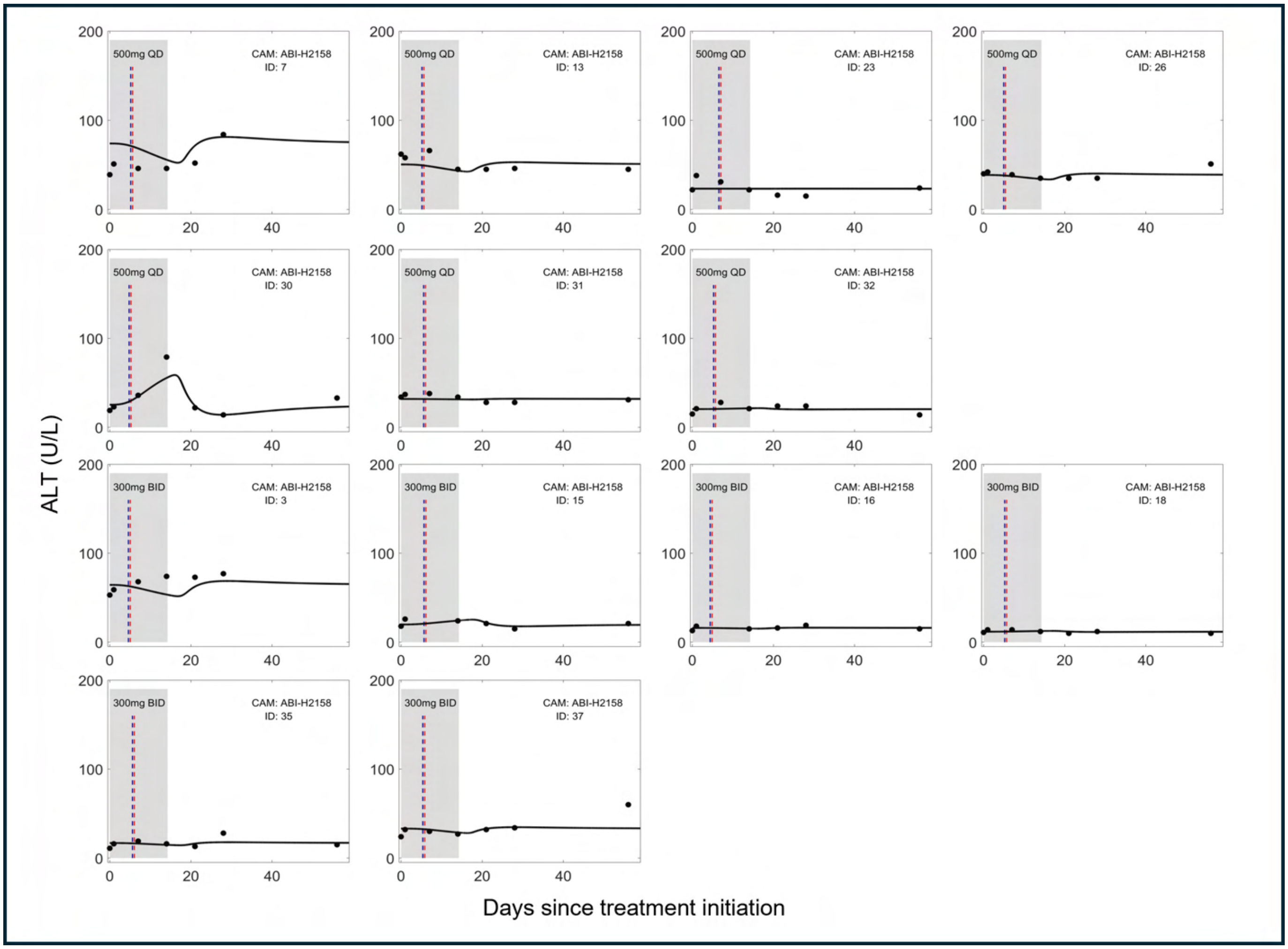


Figure S8: **The model fits to ALT data for participants in the 500 mg QD and 300 mg BID cohorts of the ABI-H2158 trial.** The ALT concentrations are shown as black circles. The model fits are shown as continuous curves. For each participant, the predicted transition time between the first and second phase of decline of HBV RNA, $t^*_{RNA}$, and HBV DNA, $t^*_{DNA}$, are shown as vertical dashed lines in blue and red, respectively.

##### Distribution of individual parameter values

We compared the estimated individual parameter values between the HBeAg-positive participants of both trials and identified a significant difference in the death rate of infected hepatocytes, $\delta$, in the individual parameter estimates ($p = 3 \times 10^{-8}$). We additionally identified a significant difference in the infection rate, $\beta$ ($p = 4 \times 10^{-8}$). As we previously showed, the infection rate $\beta$ is directly linked to the rate of viral rebound following treatment cessation [7]. Taken together, these results suggest a difference in the rate of viral rebound following treatment cessation. We recall that, rather that using a detailed pharmacokinetic model for vebicorvir or ABI-H2158, we modelled the waning antiviral effectiveness using the parameter $k$ in Eq. (S10). The population level estimates for $k$ are $k = 2.29$/day and $k = 1.5$/day, for vebicorvir and ABI-H2158, respectively, also reflect the previously observed differences in the circulating half-lives of these CAMs. We plot the distributions of the individual parameters in Fig. S9, along with the distributions of the CAM effectiveness at 100 mg or 300 mg daily dosing for both vebicorvir and ABI-H2158.

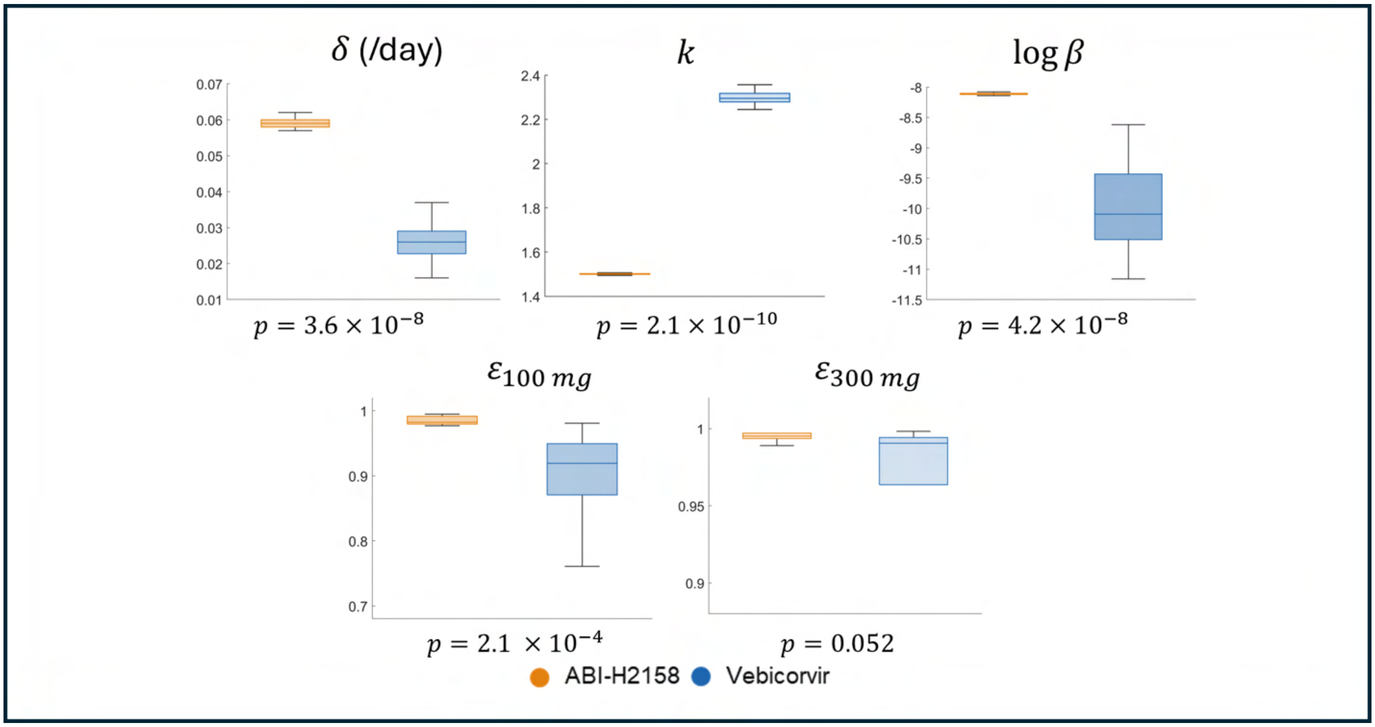


Figure S9: **The distribution of individual parameter estimates for the participants in the vebicorvir and ABI-H2158 trials.** The parameter distributions from the vebicorvir trial are shown in blue while the distributions from the ABI-H2158 trial are shown in orange. The p-value obtained from the Mann-Whitney test for each parameter distribution is shown below the plot of the distribution.

#### Validation of biomarker-effectivness relationship

In the main text, we show that the relative decline in HBV RNA and HBV DNA between baseline and day 7 directly reflects the antiviral effectiveness, expressed as $\log_{10}(1-\varepsilon)$. We validated this relationship by considering the vebicorvir and ABI-H2158 trials simultaneously. Here, we show that the predicted relationship between relative decline of these circulating biomarkers and the CAM antiviral effectiveness holds when considering these two trials individually. In Fig S10, we show the relationship between the relative decline of HBV RNA and HBV DNA between baseline and day 7 and day 14against the individual best-fit value of $\log_{10}(1-\varepsilon)$ for participants in the vebicorvir and ABI-H2158 trials individually. As in the main text, we fit a linear mixed-effects model to the antiviral effectiveness as a function of the relative HBV RNA and HBV DNA decline for each participant. We also computed the coefficient of determination for the linear mixed-effects model [9]. Consistent with our prediction, the fixed effect of this linear mixed-effects model is approximately 1 in all cases and the coefficient of determination identifies a strong relationship between the relative decline in both biomarkers and the drug effectiveness.

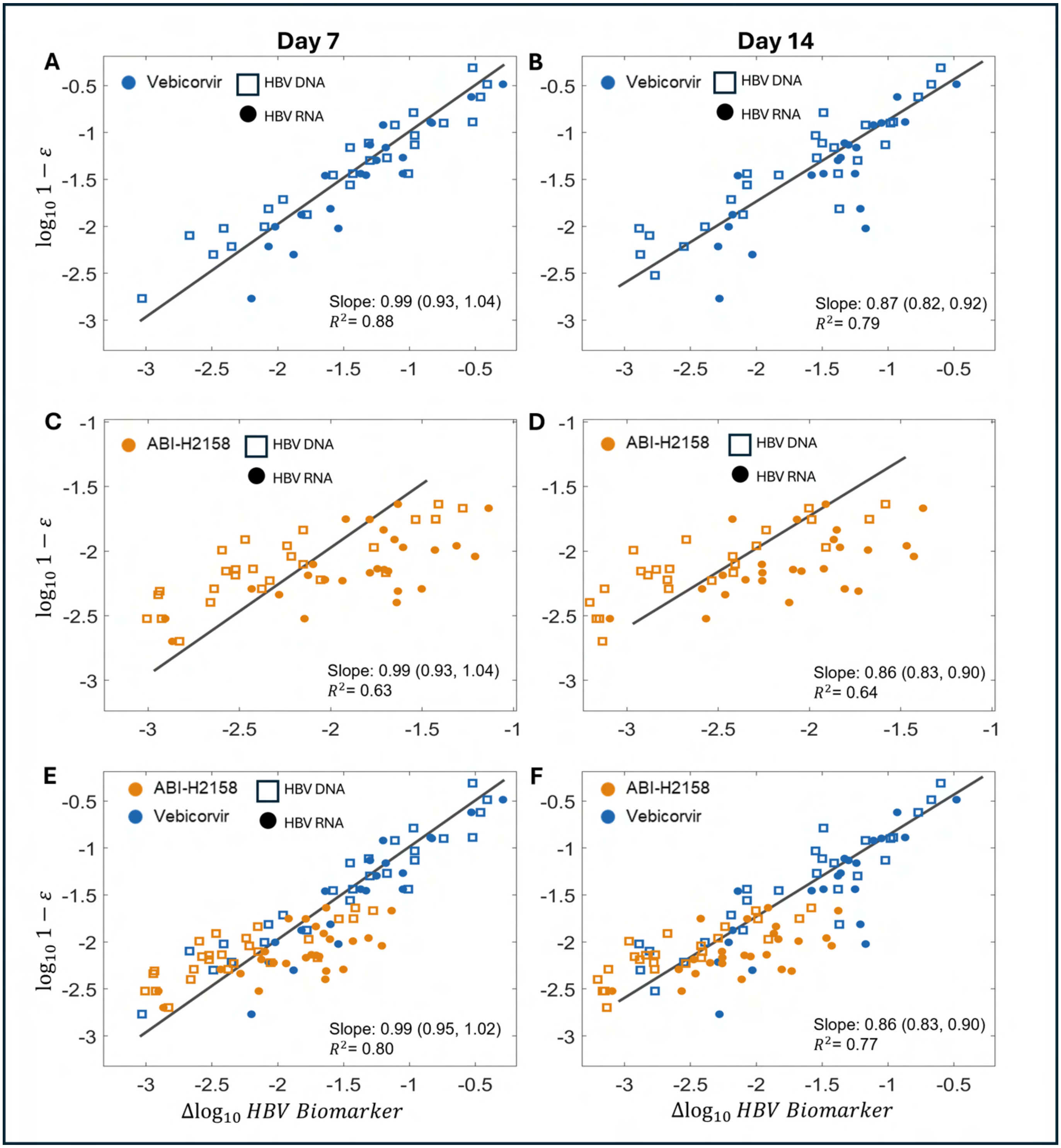


Figure S10: **Validation of Main Text Eq. (1) for the individual trials.** A scatter plot of the best-fit individual values of CAM effectiveness, $\varepsilon$, expressed as $\log_{10}(1-\varepsilon)$, against the observed relative log-decline in HBV RNA and HBV DNA in the vebicorvir and ABI-H2158 trials. The left column shows the log-decline between baseline and day 7 while the right column shows the relative decline between baseline and day 14. In all cases, the coefficient of determination, $R^2$, and the fixed effect, the slope, with 95% confidence interval of the linear mixed effects model is given in the legend.

| Trial | Baseline to Day 7<br>Grand mean (95% CI) | Deviation(95% CI) | Baseline to Day 14<br>Grand mean (95% CI) | Deviation(95% CI) |
|---|---|---|---|---|
| **Vebicorvir** | 1.02 (0.98, 1.06) | 0.09 (0.05, 0.13) | 0.91 (0.86, 0.95) | 0.10 (0.06, 0.15) |
| **ABI-H2158** | 1.00 (0.96, 1.05) | 0.11 (0.06, 0.15) | 0.88 (0.85, 1.92) | 0.09 (0.05, 0.12) |
| **Combined** | 1.01 (0.98, 1.04) | 0.10 (0.07, 0.13) | 0.89 (0.86, 0.92) | 0.09 (0.07, 0.12) |

Table S2: **Estimates for the grand mean and deviations for biomarker modulated slope mixed-effects model**. The slope of the relationship between HBV RNA and CAM effectiveness is given by the sum of the grand mean and the deviation, while the slope of the relationship between HBV DNA and CAM effectiveness is obtained by subtracting the deviation from the grand mean.

#### Biomarker modulated slope

In the linear mixed-effects model considered in the main text, we assumed that the antiviral effectiveness $\varepsilon$ was a function of the relative decline of HBV RNA and HBV DNA. Here, we test if the slope of this predictive relationship differs between biomarkers. We once again fit a linear mixed-effects model to the antiviral effectiveness as a function of the two biomarkers where we allow for the slope of this relationship to be modulated by the biomarker, either HBV RNA or HBV DNA. Once again, the predicted relationship between relative decline of these circulating biomarkers and the CAM antiviral effectiveness holds when considering this more general model. In Fig. S11, we show the relationship between the relative decline of HBV RNA and HBV DNA between baseline and day 7 or day 14 against the individual best-fit value of $\log_{10}(1-\varepsilon)$ for participants in the vebicorvir and ABI-H2158 trials individually. We report the grand mean slope of the linear mixed-effects model and coefficient of determination for the linear mixed-effects model in the legend of each panel of Fig. S11 and in Table S2 [9]. Consistent with the predicted relationship in Eq. (1) of the Main Text, 1 was contained within the 95% confidence interval for the grand mean when considering the relative decline from baseline to day 7 in both individual trials and when considering the vebicorvir and ABI-H2158 data simultaneously. As in Fig. S10, the relative decline from baseline to day 14 slightly overestimates the CAM effectiveness, which results in grand means that are slightly lower than 1.

#### Validation for the individual biomarkers

##### Relative change in HBV RNA predicts CAM effectiveness

In the main text and Fig. S12, we showed that the relative decline in both circulating biomarkers HBV RNA and HBV DNA robustly predicts $\log_{10}(1-\varepsilon)$ as a function of individuals in the vebicorvir and ABI-H2158 trials. However, the analytical result in Eq. (1) of the main text indicates that the same predictive relationship should hold between the antiviral effectiveness and the relative decline in HBV RNA. Here, we tested for this relationship in the vebicorvir and ABI-H2158 trials. Unlike in the main text, we are not considering multiple, potentially correlated, measurements from each participant. Therefore, we fit a linear regression with zero intercept to write $\log_{10}(1-\varepsilon)$ as a function of the log-decline in HBV RNA from baseline to both day 7 and day 14. In Fig. S12, we show the relationship between the log-decline in HBV RNA from baseline to day 7 in the left hand panels and from baseline to day 14 in

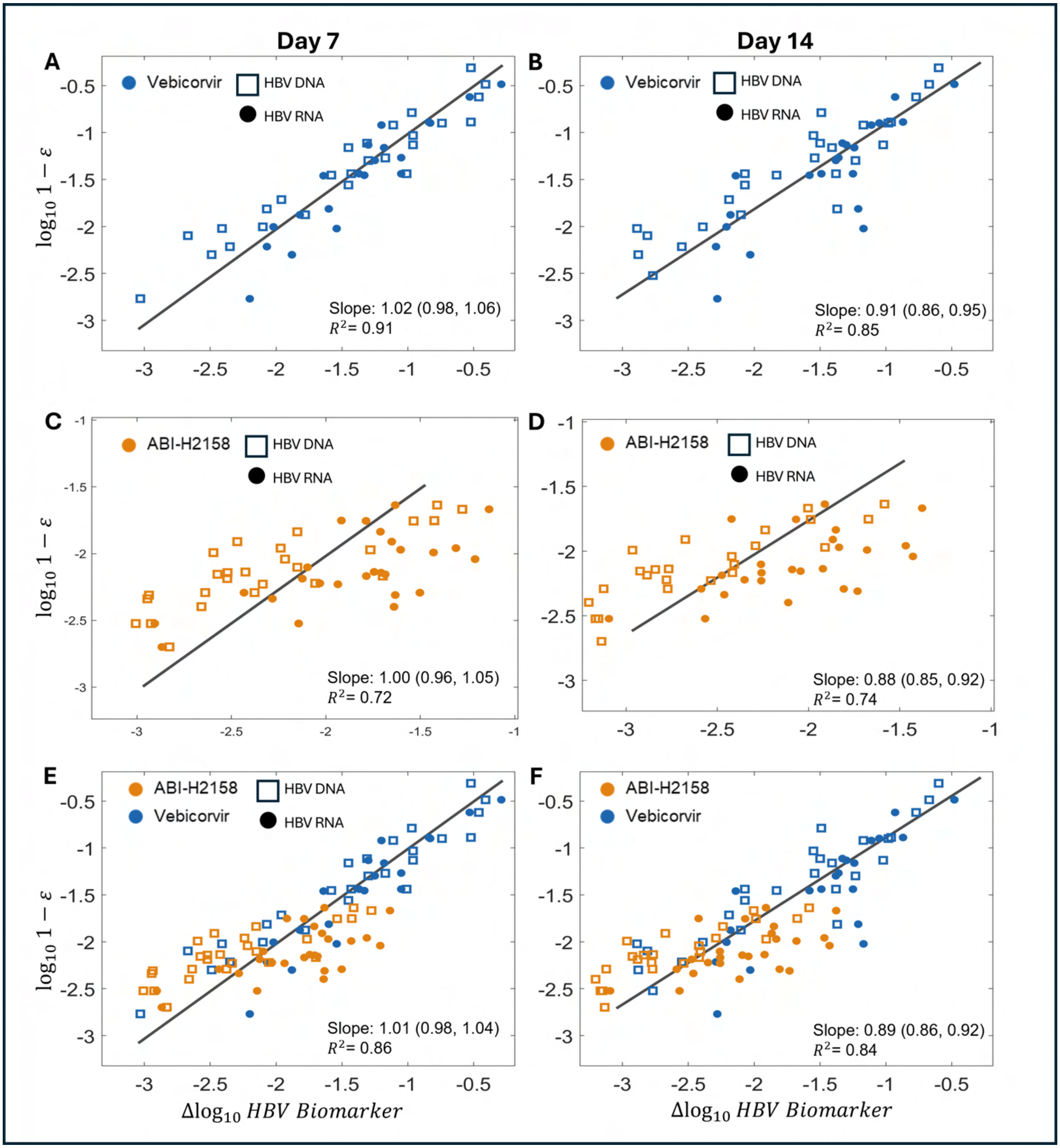


Figure S11: **Validation of Main Text Eq. (1) for the individual trials with biomarker-mediated slopes** A scatter plot of the best-fit individual values of CAM effectiveness, $\varepsilon$, expressed as $\log_{10}(1-\varepsilon)$, against the observed relative log-decline in HBV RNA and HBV DNA in the vebicorvir and ABI-H2158 trials. The left column shows the log-decline between baseline and day 7 while the right column shows the relative decline between baseline and day 14. In all cases, the coefficient of determination, $R^2$, and the fixed effect, the grand mean of the linear mixed-effects model, with 95% confidence interval, is given in the legend.

the right hand panels with the best-fit value of $\varepsilon$ for each individual participant in the respective trials. Once again, these linear regressions are consistent with the theoretical relationship between HBV RNA log-decline and CAM effectiveness in Eq. (1) of the main text. While some of the inferred slopes differ from unity, we show in the following sections that the relative decline of HBV RNA alone accurately predicts the individual antiviral effectiveness. We also show the corresponding relationship between the estimated CAM effectiveness and the log-decline in HBV RNA measured from baseline to day 7 or from baseline to day 14 for the combination of the vebicorvir and ABI-H2158 trials in the bottom row of Fig. S12.

#### Relative change in HBV DNA predicts CAM effectiveness

In the main text, we plotted $\log_{10}(1-\varepsilon)$ as a function of the relative decline in both circulating biomarkers HBV RNA and HBV DNA. However, Eq. (1) of the main text indicates that the same relationship should hold between the antiviral effectiveness and the relative decline in HBV DNA. Here, we tested for this relationship in the vebicorvir and ABI-H2158 trials. Once again, we are not considering multiple, potentially correlated, measurements from each participants. Thus, we fit a linear regression with zero intercept to write $\log_{10}(1-\varepsilon)$ as a function of the log-decline in HBV DNA from baseline to both day 7 and day 14. In Fig. S13, we show the relationship between the log-decline in HBV DNA from baseline to day 7 in the left hand panels and from baseline to day 14 in the right hand panels with the best-fit value of $\varepsilon$ for each individual participant in the respective trials. Once again, these linear regressions are consistent with the theoretical relationship between HBV DNA log-decline and CAM effectiveness in Eq. (1) of the main text. While some of the inferred slopes differ from unity, we show in the following sections that the relative decline of HBV DNA alone accurately predicts the individual antiviral effectiveness. We also show the corresponding relationship between the estimated CAM effectiveness and the log-decline in HBV DNA measured from baseline to day 7 or from baseline to day 14 for the combination of the vebicorvir and ABI-H2158 trials in the bottom row of Fig. S13.

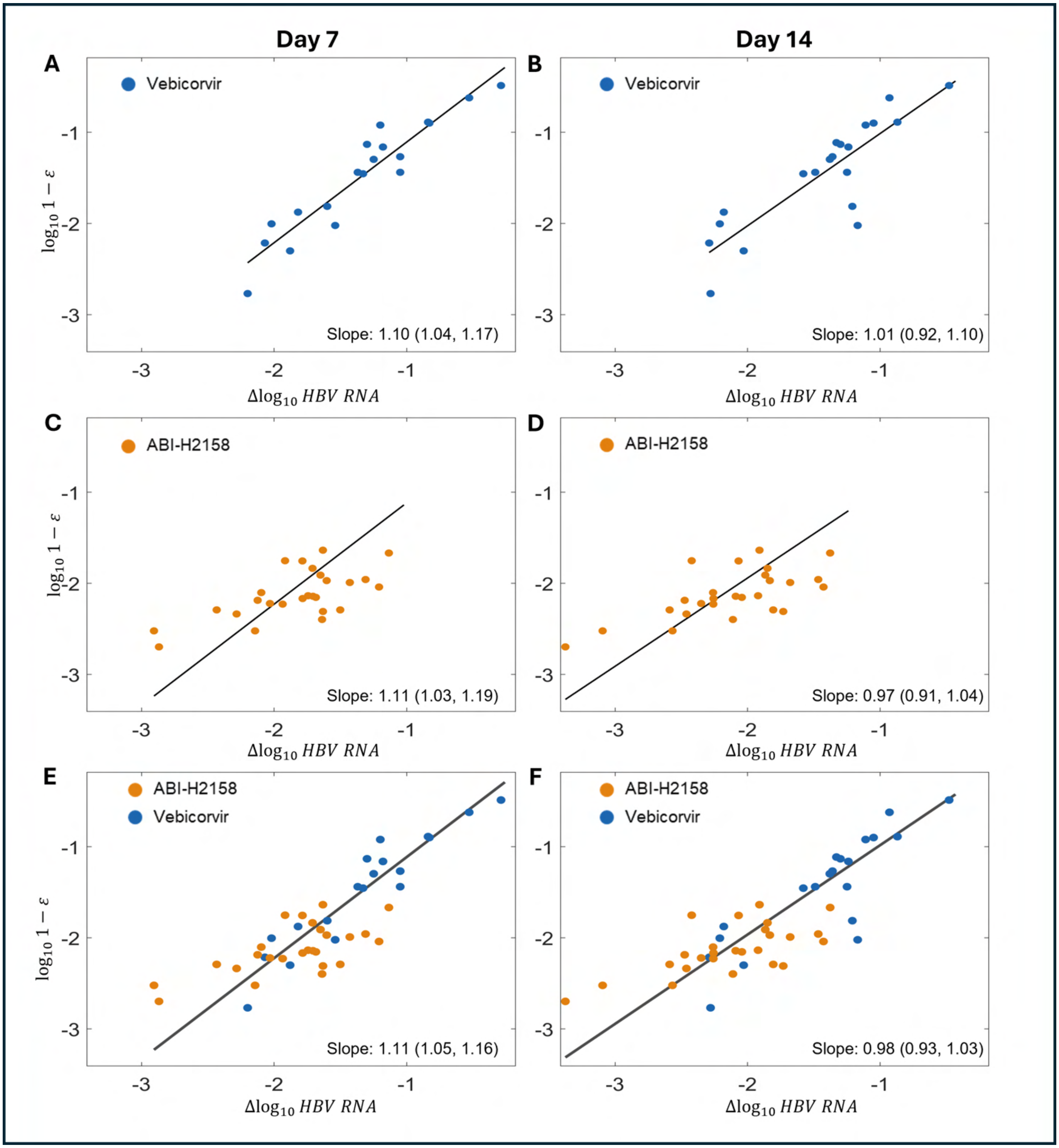


Figure S12: **The relative decline of HBV RNA directly inform CAM effectiveness for the individual trials.** Scatter plots of the best-fit CAM effectiveness, expressed as $\log(1-\varepsilon)$, against the observed relative log HBV RNA decline. Plots A and B show the relative HBV RNA decline at day 7 and day 14 for each individual participant in the vebicorvir trial. Plots C and D show the relative HBV RNA decline at day 7 and day 14 for each individual participant in the ABI-H2158 trial, respectively. Finally, plots E and F show the scatter plot of the best-fit CAM effectiveness against the relative log-decline in HBV RNA measured on day 7 and day 14 respectively, for the combined trials. Each legend reports the slope and 95% confidence interval of the line of best fit.

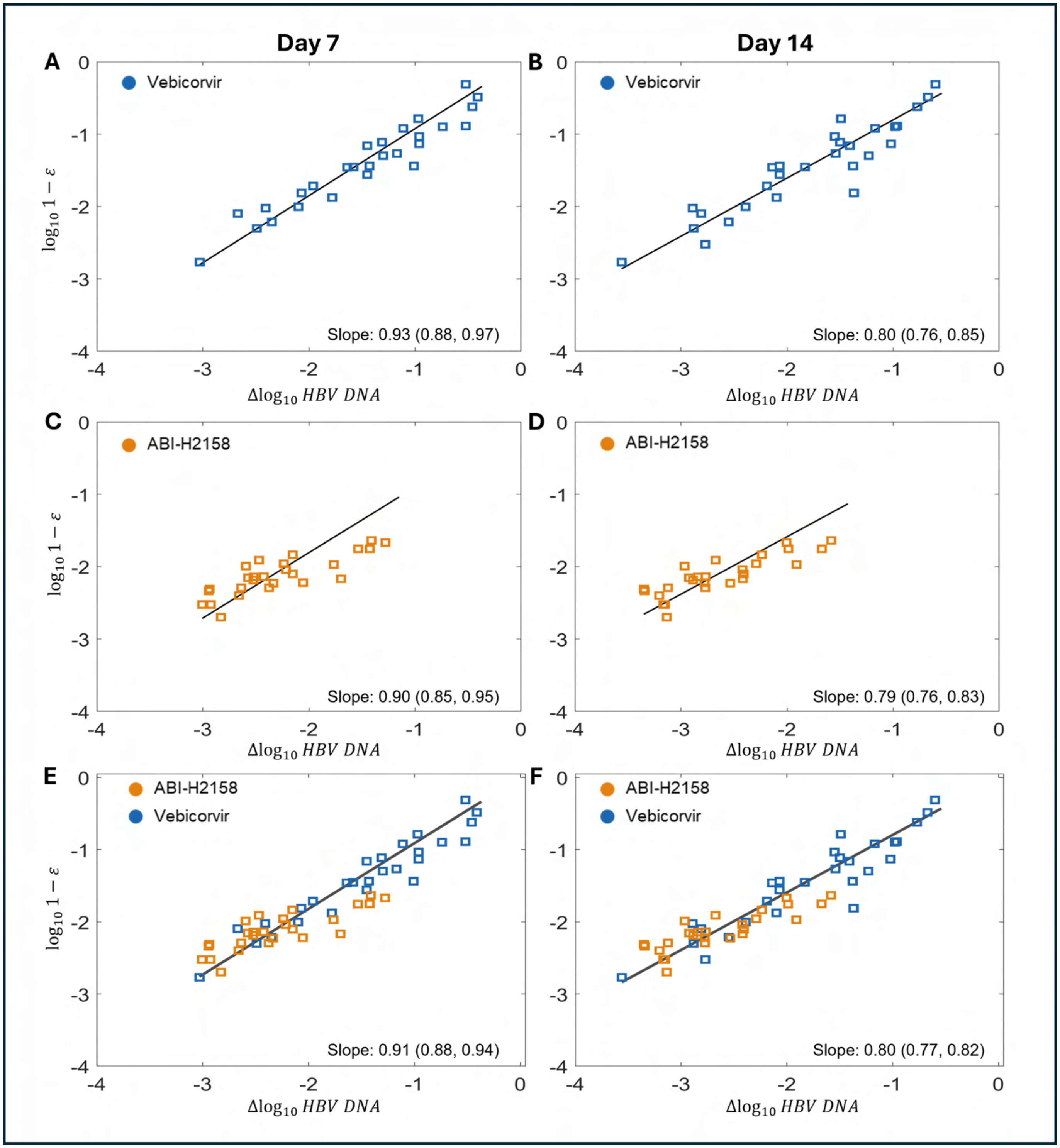


Figure S13: **The relative decline of HBV DNA directly inform CAM effectiveness for the individual trials.** Scatter plots of the best-fit CAM effectiveness, expressed as $\log(1 - \varepsilon)$, against the observed relative log HBV DNA decline. Plots A and B show the relative HBV DNA decline at day 7 and day 14 for each individual participant in the vebicorvir trial. Plots C and D show the relative HBV DNA decline at day 7 and day 14 for each individual participant in the ABI-H2158 trial, respectively. Finally, plot E and F show the scatter plot of the best-fit CAM effectiveness against the relative log-decline in HBV DNA measured on day 7 and day 14 respectively, for the combined trials. Each legend reports the slope of the line of best fit and 95% confidence interval.

#### Relative error of predicting individual antiviral effectiveness

##### Predicting antiviral effectiveness using both HBV RNA and HBV DNA

As shown in Eq. (2) of the main text, we can directly compare the best-fit effectiveness, $\varepsilon$, against the relative decline in HBV RNA and HBV DNA for each participant. However, we may not have an observation of HBV RNA or HBV DNA made at $t^*_{RNA}$ or $t^*_{DNA}$ in practice. This may lead to some deviations from the predicted relationship between $\varepsilon$ in Eq. (2) of the main text. We therefore use the relative decline of both these biomarkers to estimate the antiviral effectiveness for the $i-$th participant via

$$\hat{\epsilon}_{7,RNA,i} = 1 - R_i(7)/R_i(0)) \quad \text{and} \quad \hat{\epsilon}_{14,RNA,i} = 1 - R_i(14)/R_i(0)),$$

and

$$\hat{\epsilon}_{7,DNA,i} = 1 - V_i(7)/V_i(0)) \quad \text{and} \quad \hat{\epsilon}_{14,DNA,i} = 1 - V_i(14)/V_i(0)).$$

We then estimate the individual antiviral effectiveness by

$$\hat{\epsilon}_{7,i} = \frac{\hat{\epsilon}_{7,DNA,i} + \hat{\epsilon}_{7,RNA,i}}{2}, \quad \text{and} \quad \hat{\epsilon}_{14,i} = \frac{\hat{\epsilon}_{14,DNA,i} + \hat{\epsilon}_{14,RNA,i}}{2},$$

i.e., the mean of these two estimates at day 7 or day 14. We calculate the relative error between the best-fit estimate for $\varepsilon_i$ and $\hat{\epsilon}_{7,i}$ or $\hat{\epsilon}_{14,i}$ via

$$\text{Relative error}_{7,i} = 100\left(\frac{\varepsilon_i - \hat{\epsilon}_{7,i}}{\varepsilon_i}\right) \quad \text{and} \quad \text{Relative error}_{14,i} = 100\left(\frac{\varepsilon_i - \hat{\epsilon}_{14,i}}{\varepsilon_i}\right)$$

This percent relative error measures the accuracy of the predicted effectiveness for each individual participant, and can be either positive or negative, depending if $\hat{\epsilon}$ under- or overestimates the best-fit effectiveness, respectively.

We show the distribution of relative errors for each participant in the vebicorvir and ABI-H2158 trials for $\hat{\epsilon}$ in Fig. S14. In nearly all cases, the estimated effectiveness $\hat{\epsilon}$ is a good approximation of the true fit value, $\varepsilon$ as illustrated by the clustering of relative errors near 0% and the median error being below 1.1% in all cases.

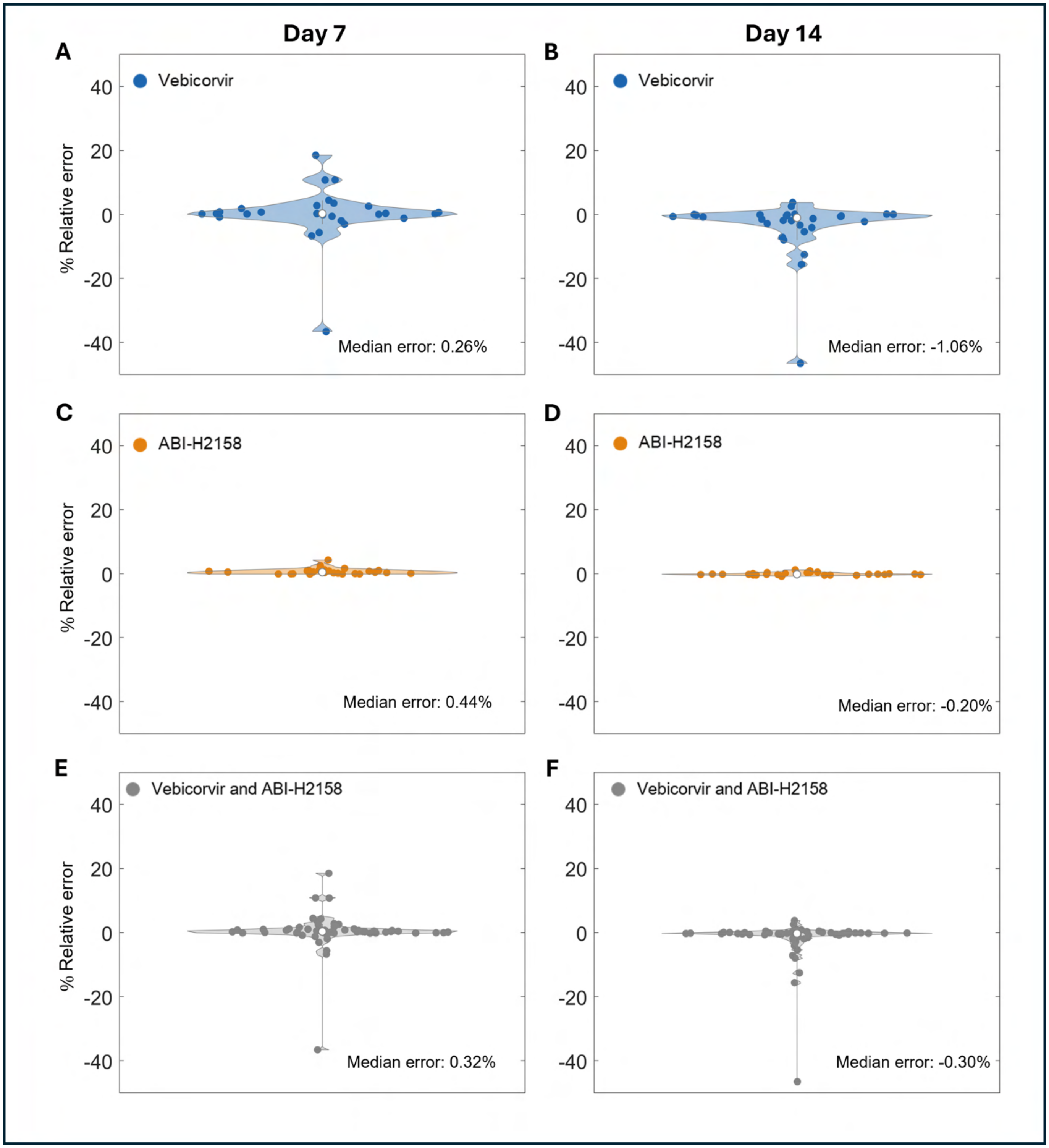


Figure S14: **The relative error of predicting antiviral effectiveness using both HBV RNA and HBV DNA** Violin plots of the relative error between the best-fit CAM effectiveness, $\varepsilon$, against the effectiveness predicted based on the observed HBV RNA and HBV DNA decline at day 7 and day 14, $\hat{\epsilon}$, for the individual trials. The left column shows the relative error obtained between the best-fit value of $\varepsilon$ and the estimated value of $\hat{\epsilon}$ from the observed decline of HBV RNA and HBV DNA between baseline and day 7. The right column shows the corresponding percent relative error for decline between baseline and day 14. Panels (A) and (B) show the results from the vebicorvir trial, (C) and (D) from the ABI-H2158 trial, and (E) and (F) from the combined trials. Each legend reports the median relative error between the fit and estimated effectiveness, which is shown as a white dot in each panel.

#### Predicting antiviral effectiveness using HBV RNA only

As before, we use the relative decline in HBV RNA to predict the CAM effectiveness for the individual participants. Specifically, we set

$$\hat{\epsilon}_{7,i} = 1 - R_i(7)/R_i(0)) \quad \text{and} \quad \hat{\epsilon}_{14,i} = 1 - R_i(14)/R_i(0)).$$

We then calculate the relative error between the best-fit estimate for $\varepsilon_i$ and the relative HBV RNA decline at day 7 or day 14 via

$$\text{Relative error}_{7,i} = 100 \left( \frac{\varepsilon_i - \hat{\epsilon}_{7,i}}{\varepsilon_i} \right) \quad \text{and} \quad \text{Relative error}_{14,i} = 100 \left( \frac{\varepsilon_i - \hat{\epsilon}_{14,i}}{\varepsilon_i} \right)$$

We show the distribution of relative errors for each participant in the vebicorvir and ABI-H2158 trials in Fig. S15. Once again, the estimated effectiveness $\hat{\epsilon}$ is a good approximation of the true fit value, $\varepsilon$, as illustrated by the clustering of relative errors near 0% and the median error being below 1% in all cases, which further validates our predictive relationship between the CAM effectiveness and the relative log decline in HBV RNA.

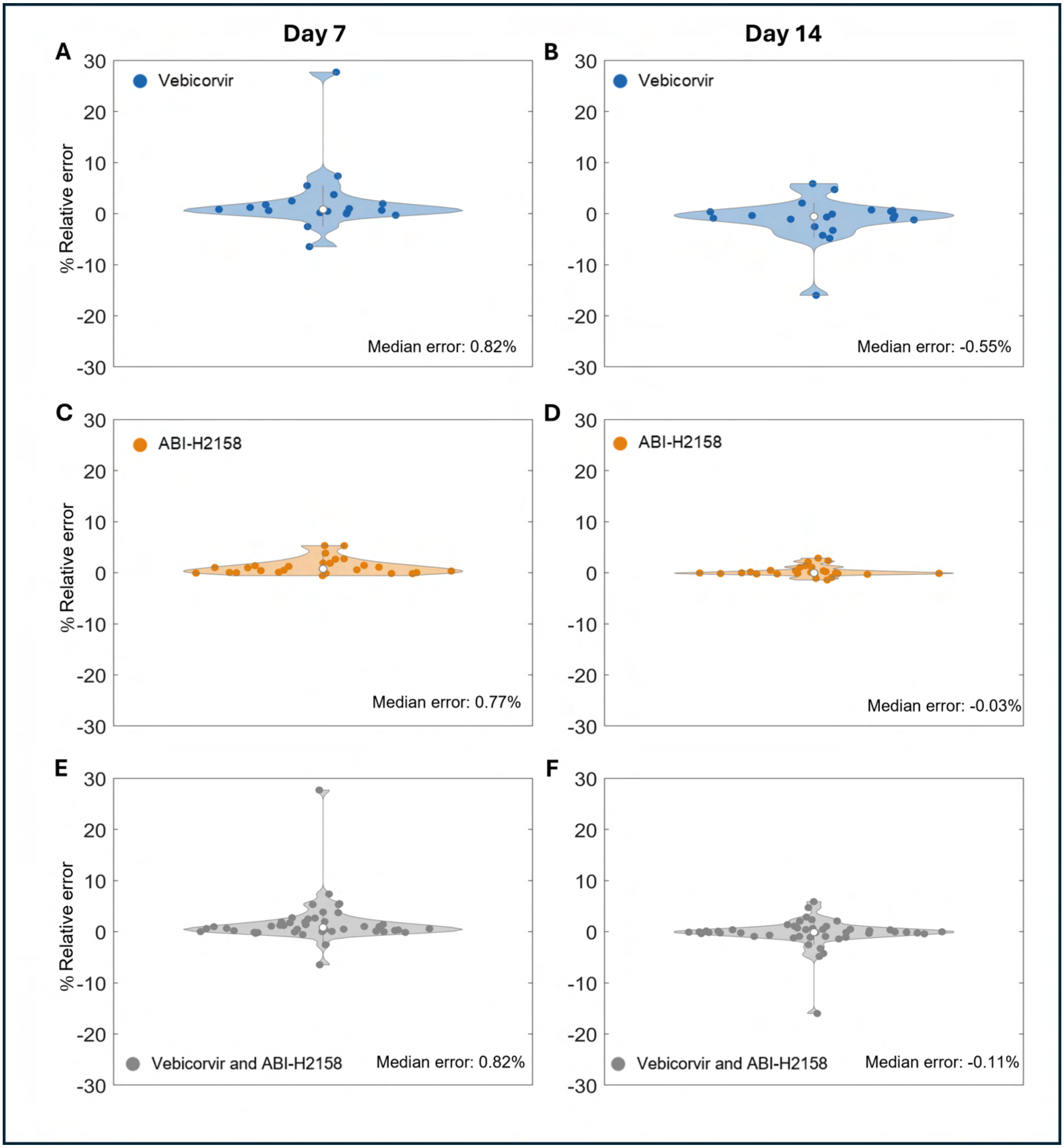


Figure S15: **The relative error of predicting antiviral effectiveness using HBV RNA only.** Violin plots of the relative error between the best-fit CAM effectiveness, $\varepsilon$, against the effectiveness predicted based on the observed HBV RNA decline at day 7 and day 14, $\hat{\varepsilon}$. The left column shows the relative error obtained between the best-fit value of $\varepsilon$ and the estimated value of $\hat{\varepsilon}$ from the observed decline of HBV RNA between baseline and day 7. The right column shows the corresponding percent relative error for decline between baseline and day 14. Panels (A) and (B) show the results from the vebicorvir trial, (C) and (D) from the ABI-H2158 trial, and (E) and (F) from the combined trials. Each legend reports the median magnitude of the relative error and the median relative error between the fit and estimated effectiveness, which is shown as a white dot in each panel.

##### Predicting antiviral effectiveness using HBV DNA only

As we did for HBV RNA, we use the relative decline in HBV DNA to predict the CAM effectiveness for the individual participants. Specifically, we set

$$\hat{\epsilon}_{7,i} = 1 - V_i(7)/V_i(0)) \quad \text{and} \quad \hat{\epsilon}_{14,i} = 1 - V_i(14)/V_i(0)).$$

We then calculate the relative error between the best-fit estimate for $\varepsilon_i$ and the relative HBV DNA decline at day 7 or day 14 via

$$\text{Relative error}_{7,i} = 100\left(\frac{\varepsilon_i - \hat{\epsilon}_{7,i}}{\varepsilon_i}\right) \quad \text{and} \quad \text{Relative error}_{14,i} = 100\left(\frac{\varepsilon_i - \hat{\epsilon}_{14,i}}{\varepsilon_i}\right)$$

We show the distribution of relative errors for each participant in the vebicorvir and ABI-H2158 trials in Fig. S16. Once again, the estimated effectiveness $\hat{\epsilon}$ is a good approximation of the true fit value, $\varepsilon$, as illustrated by the clustering of relative errors near 0% and the median error being below 2.1% in all cases, which further validates our predictive relationship between the CAM effectiveness and the relative log decline in HBV DNA.

#### Clinical trial simulations

We generated virtual populations by sampling from the posterior population parameter distributions obtained by fitting the clinical data from the ABI-H2158 trial. The distributions of all parameters, except $\varepsilon$, were independent of CAM effectiveness. As is commonly done [3, 10], we sampled 5000 parameter sets from the posterior parameter distributions to represent 5000 virtual trial participants. We incorporated the antiviral effects of our hypothetical CAMs by changing the mean population estimate for $\varepsilon$, but not the variance of the population distribution of $\varepsilon$, to correspond to the hypothetical CAM. We simulated clinical trials by simulating the viral dynamics model in Eq. (S1) for each of the 5000 individual parameter sets and measuring the simulated HBV RNA and HBV DNA.

In the main text, we considered a hypothetical next-generation class-E CAM with mean population effectiveness $\bar{\varepsilon} = 0.9999$. This hypothetical CAM is 50-fold more effective than the 300 mg QD dose of ABI-H2158 and 160-fold more effective than the 300 mg QD dose of vebicorvir. It is important to note that a 4th generation CAM, ABI-4334, is 500-fold more effective in than vebicorvir as measured in *in vitro* dose-response assays [14], which implies that the high effectiveness of our hypothetical CAM may be observed with next-generation CAMs currently undergoing clinical trials.

We wished to test the robustness of our method to determine CAM effectiveness to the timing of the second measurement of HBV RNA and HBV DNA. We compared the most effective dose, 300 mg QD, of ABI-H2158 against the hypothetical CAM in 10,000 virtual clinical trials, each with 10 virtual participants in each treatment arm (20 participants total per trial). In each of the 10,000 virtual clinical trials, we calculated the log-decline in HBV RNA

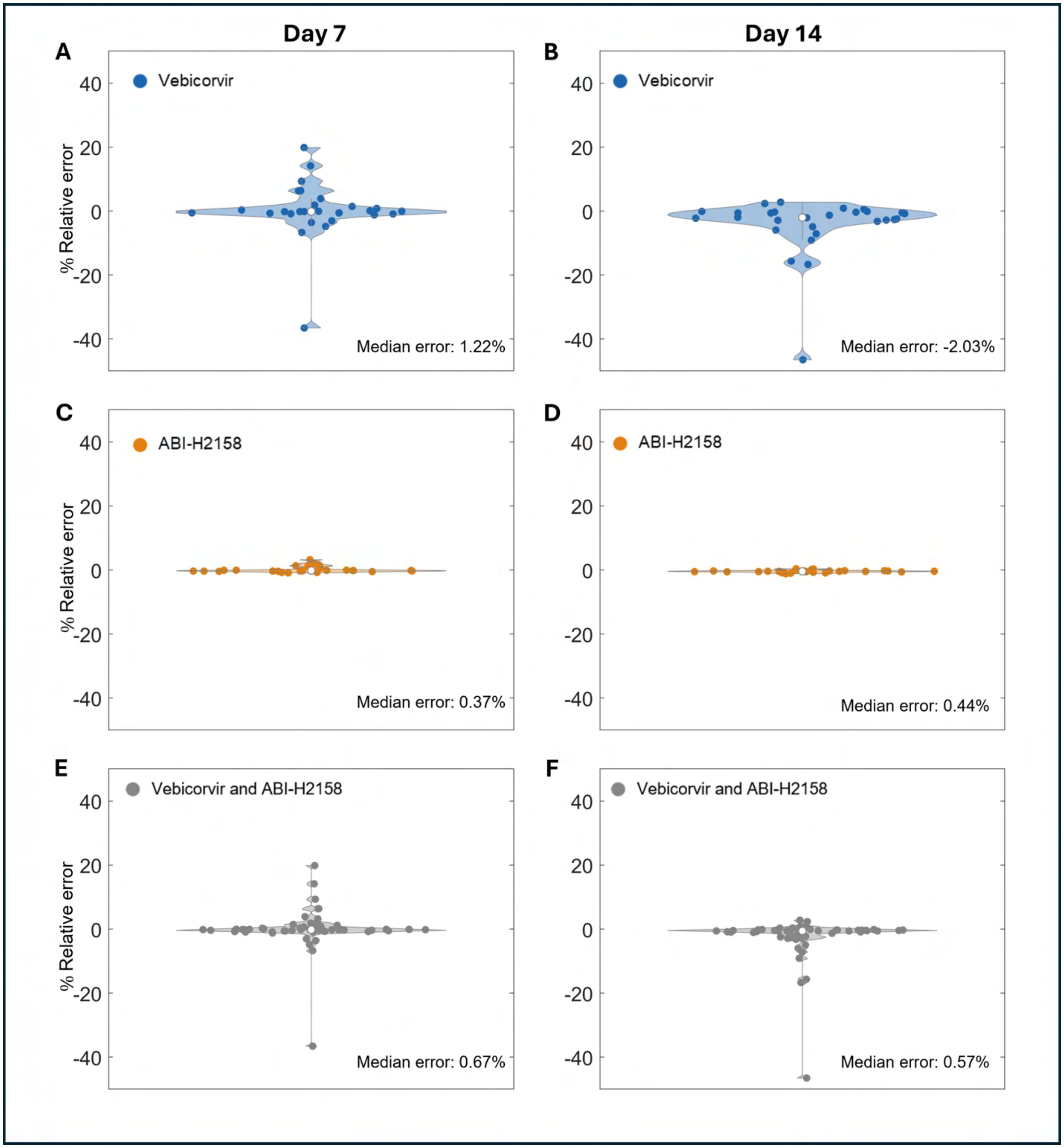


Figure S16: **The relative error of predicting antiviral effectiveness using HBV DNA only.** Violin plots of the relative error between the best-fit CAM effectiveness, $\varepsilon$, against the effectiveness predicted based on the observed HBV DNA decline at day 7 and day 14. The left column shows the relative error obtained between the best-fit value of $\varepsilon$ and the estimated value of $\hat{\epsilon}$ from the observed decline of HBV DNA between baseline and day 7. The right column shows the corresponding percent relative error for decline between baseline and day 14. Panels (A) and (B) show the results from the vebicorvir trial, (C) and (D) from the ABI-H2158 trial, and (E) and (F) from the combined trials. Each legend reports the median magnitude of the relative error and the median relative error between the fit and estimated effectiveness, which is shown as a white dot in each panel.

and HBV DNA between baseline and day 14 for each virtual participant and used these values to calculate both the individual and mean population effectiveness $\varepsilon$ using Main Text Eq. (1). We tested our method for calculating CAM effectiveness for participants in a randomly selected virtual clinical trial, and plot the simulated day 14 relative decline in HBV RNA and HBV DNA against the known value of $\log_{10}(1-\varepsilon)$ for the 20 virtual participants in this trial in Fig. S17A. We next compared the distributions of the predicted mean CAM effectiveness for the hypothetical CAM and 300mg QD of ABI-H2158 obtained from Main Text Eq. (1) against the known population efficacies in Fig. S17B. The population effectiveness of both ABI-H2158 and the hypothetical CAM are well-estimated using the relative decline in HBV RNA and HBV DNA at day 14, and the 95% confidence interval of the mean population estimates clearly differentiates 300 mg QD of ABI-H2158 from the hypothetical CAM.

We also tested if the relative decline in HBV RNA and HBV DNA at day 10 would robustly predict the antiviral effectiveness of the hypothetical CAM. However, the first phase of decline of both HBV RNA and HBV DNA extends past day 10 for this hypothetical CAM. Consequently, the relative HBV RNA and HBV DNA decline on day 10 both significantly under-estimate the population effectiveness (not shown). Indeed, more effective CAMs lengthen the duration of the first phase of decline, i.e. $t^*_{RNA}$ and $t^*_{DNA}$ are increasing functions of CAM effectiveness. Consequently, we suggest using the relative decline between baseline and day 14 to ensure that the HBV RNA and HBV DNA dynamics capture the full CAM mediated effect during the first phase of decline.

Using our virtual clinical trial approach, we next tested how large of a cohort would be needed to accurately estimate the population effectiveness by calculating the relative log-decline at day 14. For cohort sizes of $N = 5, 6, 7, ..., 25$, we performed 10,000 bootstrapped simulated clinical trials. In Panel A of Fig. S18, we show the comparison between the estimated mean population effectiveness, calculated via the log-decline in HBV RNA and HBV DNA at day 14, against the true effectiveness $\bar{\varepsilon}$ for trial cohorts of different sizes. For cohort sizes of 5-25 participants, we note that the true value of $\bar{\varepsilon}$ falls within the 95% confidence interval of the mean, although the 95% confidence intervals are relatively large for cohorts of less than 10 participants. We therefore conclude that 10 participants is a sufficiently large cohort to accurately estimate the true CAM effectiveness using our method. However, we note that the relative decline of HBV DNA consistently overestimates the true effectiveness of the hypothetical CAM for larger cohort sizes. Nevertheless, it is important to note that our method for calculating the antiviral effectiveness of the CAM results in a relative error of approximately $1 \times 10^{-3}$%, as the approximation is correct to the 5th decimal place.

Recent experimental evidence has indicated that class-A CAMs, such as RG7907, which induce the formation of abherrent capsids, rather than the empty capsids that occur during treatment with class-E CAMs such as vebicorvir and ABI-H2158, may increase the death rate of HBV infected hepatocytes. However, our conclusion that the log-decline of HBV RNA and HBV DNA at day 14 is a robust indicator of CAM effectiveness relies on balancing the error induced by measuring the log-decline before the completion of the first phase of decline against the error induced by death of infected hepatocytes during the second phase of decline. Consequently, the increased death rate of infected hepatocytes during treatment with a class-A CAM may lead to increased decline of HBV RNA and HBV DNA between $t^*_{RNA}$ (or $t^*_{DNA}$) and day 14 and thus result in overestimates of CAM effectiveness. We therefore considered a hypothetical class-A CAM with effectiveness $\bar{\varepsilon} = 0.9999$ and a secondary mechanism of action that doubles the death rate of infected hepatocytes, $\delta$, during treatment. We generated a virtual population as before with a population estimate for $\delta = 0.11$/day. We note that this population estimate for $\delta$ is in-line with estimates for this parameter from the modelling of RG7907, a class-A CAM, by Gonçalves et al. [5]. Once again, we simulated daily treatment with this hypothetical class-A CAM in 5000 virtual patients. For cohort sizes of $N = 5, 6, 7, ..., 30$, we completed 10,000 bootstrapped clinical trials and estimated the mean effectiveness by calculating the log-decline in HBV RNA and HBV DNA at day 14 (Fig. S18B). The true effectiveness is within the 95% confidence interval for all values of $N = 5, 6, 7, ...30$, although the 95% confidence intervals are once again large for cohorts of less than 10 participants, and the true effectiveness is near the upper limit of the confidence interval for cohorts larger than 23 participants. This indicates that the log-decline in HBV RNA or HBV DNA at day 14 both robustly predicts the effectiveness of this hypothetical class-A CAM for clinically reasonable cohort sizes. Once again, the relative decline of HBV DNA consistently overestimates the true effectiveness of this hypothetical class-A CAM for larger cohort sizes. However, it is important to note that our method for calculating the antiviral effectiveness of the CAM results in a relative error of approximately $5 \times 10^{-3}$%.

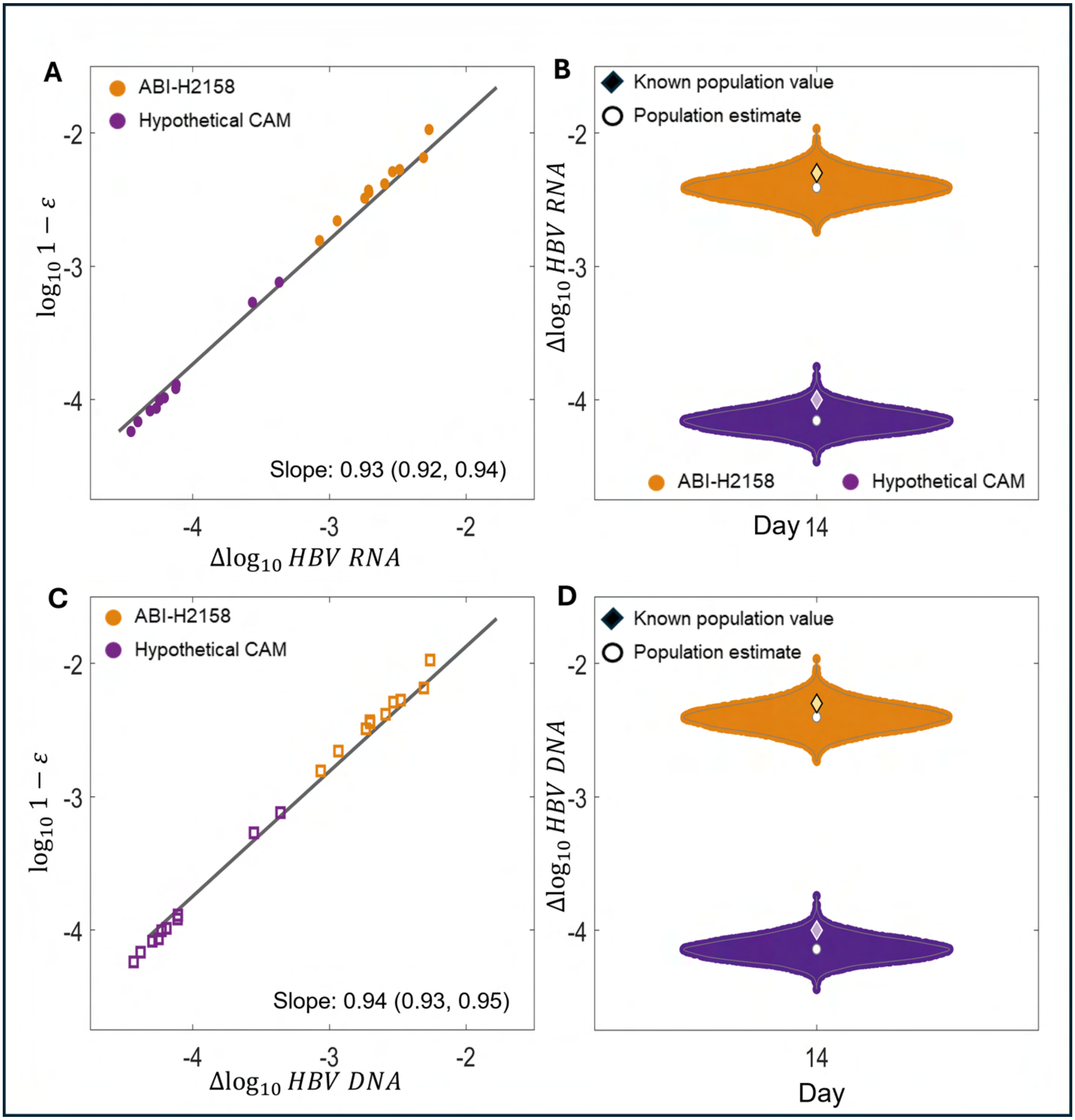


Figure S17: **Viral kinetics at day 14 robustly predicts CAM effectiveness and distinguishes between next-generation CAMs.** (A) A scatter plot of the known values of CAM effectiveness, $\varepsilon$, plotted as $\log(1-\varepsilon)$, against the simulated relative log-decline in HBV RNA at day 14 in a randomly selected cohort of 10 virtual patients in simulated clinical trials of the hypothetical CAM and 300 mg QD of ABI-H2158. (B) Violin plots of 10,000 estimates for the mean population effectiveness of the hypothetical CAM and 300 mg QD ABI-H2158 using clinical trials with 10 participants. (C) A scatter plot of the known values of CAM effectiveness, $\varepsilon$, plotted as $\log(1-\varepsilon)$, against the simulated relative log-decline in HBV DNA at day 14 in the same cohort of 10 virtual patients as (B). (D) Violin plots of 10,000 estimates for the mean population effectiveness of the hypothetical CAM and 300 mg QD ABI-H2158 using clinical trials with 10 participants. In (B) and (D), the population effectiveness is calculated by the relative HBV RNA or HBV DNA, respectively, log-decline between baseline and day 14 in each trial. The known population estimates are shown as diamonds while the estimated values are shown as white circles. In all cases, the relative error of the population estimate is less than 0.11%.

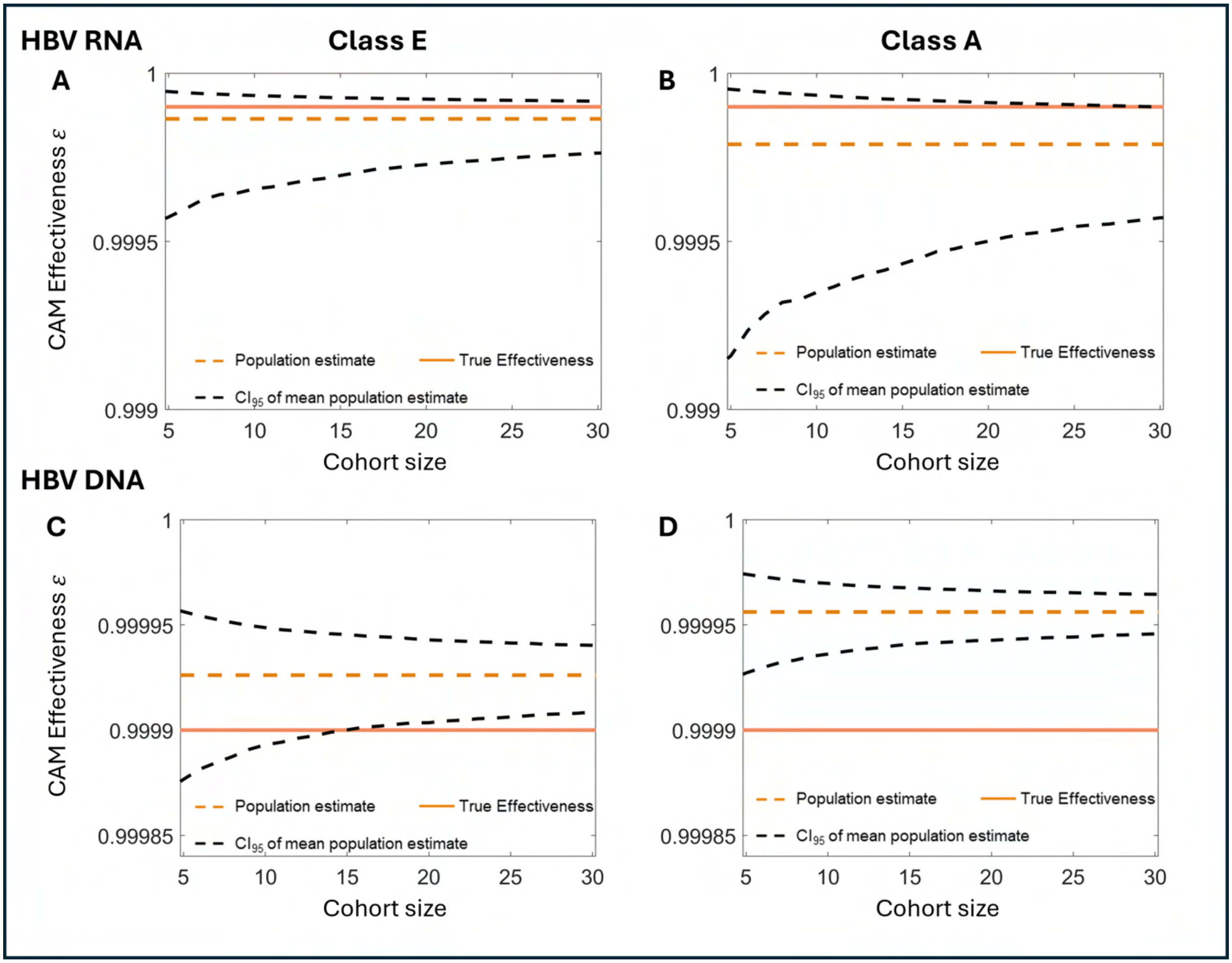


Figure S18: **Simulated clinical trial results for hypothetical class E and class A CAMs.** (A) The true CAM effectiveness against the estimated effectiveness calculated by the relative HBV RNA log-decline at day 14 in a clinical trial of the hypothetical class-E CAM hypothetical as a function of the number of participants. (B) The true CAM effectiveness against the estimated effectiveness calculated by the relative HBV RNA log-decline at day 14 in a clinical trial of the hypothetical class-A CAM as a function of the number of participants. (C) The true CAM effectiveness against the estimated effectiveness calculated by the relative HBV DNA log-decline at day 14 in a clinical trial of the hypothetical class-E CAM hypothetical as a function of the number of participants. (D) The true CAM effectiveness against the estimated effectiveness calculated by the relative HBV DNA log-decline at day 14 in a clinical trial of the hypothetical class-A CAM as a function of the number of participants. In all cases, the true CAM effectiveness is in solid orange, while the population estimate from the log-decline in HBV RNA at day 14 is in dashed orange. The dashed black lines show the 95% confidence interval of the mean calculated by 10,000 bootstrap samples for each trial size. While these estimates lie outside the 95% CI, the relative error is roughly 0.005%.
